# The micro-hotspots of cholera in Kano State, Nigeria, 2010-2019—analysis of patient characteristics, Spatio-temporal patterns and contextual determinants at the ward level

**DOI:** 10.1101/2021.08.20.21262313

**Authors:** Moise Chi Ngwa, Chikwe Ihekweazu, Tochi Joy Okwor, Sebastian Yennan, Nanpring Williams, Kelly Elimian, Nura Yahaya Karaye, James Agada Oche, Imam Wada Bello, David A. Sack

## Abstract

Cholera is endemic in Nigeria, and Kano State reports outbreaks yearly with a case fatality rate (CFR) of 3.3% from 2010 to 2019. The lack of data at ward level has enabled the disease to evade focused interventions. The goal of this study was to describe the geographic distributions, care-seeking behaviors, Spatio-temporal cluster patterns of the micro-hotspots (hotspots wards) linked with suspected and confirmed cases and deaths of cholera in Kano State.

Suspected and confirmed cholera morbidity and mortality at the ward level from 2010-2019 were acquired from the Nigeria Centre for Disease Control. Population and waterbody data were obtained from the Nigeria Expanded Program on Immunization and online, respectively. Data analysis used SaTScan and methods recommended by the Global Task Force on Cholera Control.

During these ten years, 18,483 suspected and confirmed cases (617 deaths) were reported with 67.7% of the cases and 72% of the deaths from rural wards. The ages of the cases ranged from 1 month to 100 years with a distribution skewed to the older years. CFRs were statistically higher in the <5-year olds compared to those >14 years (*p-value* = 0.0005). For 2010-2019, gender was statistically associated with cholera outcome (survived/died) (*p-value* = 0.0006), and women in the rural setting disproportionately died from cholera than women in the urban area (*p-value* = 0.003). Cholera severity, as measured by hospitalization and death, was higher in the urban (77.4%) compared with the rural (53.4%) setting with the highest severity (84.7%) registered among those >14 years. Rapid Diagnostic Tests (RDT) were performed in 1.3% (249) samples of all suspected cases and ranged from 0.7% among the 5-14 year-olds in the rural to 3.5 % among the < 5-year-olds in the urban areas. Of the stool samples collected, 62.7% tested positive for *V. cholerae* using RDT. The positivity rate was least in the urban setting amongst the <5 years (41.2%) while care-seeking-behavior ranged from 52.1% in the urban to 82.7% in the rural settings. Seasonal patterns of disease often differed between urban and rural settings with outbreaks occurring in both the dry and rainy seasons, but with more intense transmission occurring during the rainy season from week 22 (early June) to week 40 (late September). A Spatio-temporal clustering analysis detected 168 micro-hotspots out of 404 wards, with a population of 4,876,254, having a significantly higher risk (relative risk 1.01-18.73) compared to the State as a whole. While 79 micro-hotspots with a population of 2,119,974 had a RR ≥ 2. The micro-hotspots tended to cluster around waterbodies. SaTScan and GTFCC methods generally agreed in micro-hotspots detection.

This study shows the epidemiology of cholera in Kano State differs between urban and rural settings and that hotspot maps at the ward level, not hotpots maps at the Local Government Area level, are best suited for targeting interventions including vaccines. Appropriate studies are needed to further delineate the urban and rural divide of outbreaks but targeting interventions to the identified high-priority micro-hotspots will facilitate cholera elimination from the state.

**Author summary:** Cholera is endemic in Nigeria since 1970, and Kano State reports outbreaks almost every year. From 2010-2019, Kano State reported 18,483 cases and 617 deaths, for a case fatality rate of 3.3%. Focusing interventions at the Local Government Area instead of the ward (local) level contributed to the continuous threat from cholera in Kano State. When we divided the state into its two geographic areas (urban and rural), there were very different epidemiology as well as Spatio-temporal patterns of clustering of wards with elevated relative risk (micro-hotspots). Nearly two-thirds of the cases and deaths were reported from rural wards. The ages of the cases ranged from 1 month to 100 years with case fatality ratios higher in the <5-years olds compared to those >14 years (*p-value* = 0.0005). Women in the rural area not only had more cases but also were more likely to die from cholera than women in the urban area (*p-value* = 0.003). The hospitalization rate was higher in the urban than rural setting whereas care-seeking behavior was higher in rural than urban areas. Rapid Diagnostic Tests to confirm cholera was very low overall. Seasonal patterns of disease differed between urban and rural areas with outbreaks occurring in both the dry and rainy seasons, but with more cases from June to September, during the rainy season. A population of 4,876,254 live in the 168 micro-hotspots in which cholera risks were 1.01 to 18.73 times higher compared to the State as a whole. Following the Global Task Force on Cholera Control recommendations, interventions should focus on these identified micro-hotspots for cholera elimination from Kano State.

## Introduction

Cholera is a waterborne infection that makes over a million people sick yearly with an estimated 95,000 deaths worldwide [1]. In the last decade, Sub-Saharan Africa (SSA) reported the greatest cases and deaths with the Democratic Republic of Congo and Nigeria contributing the greatest burden of cases in Africa. Nigeria first reported cholera in 1970 [2, 3] and this marked the beginning of an endemic pattern characterized by 334,990 reported cases with a case fatality ratio (CFR) of 5.8% between 1991 and 2019 [2, 4]. While reported cases decreased from 45,037 in 2018 to 2,486 in 2019 [5, 6], the decrease cannot be attributed to improved interventions, but rather to cholera’s pattern of episodic cycles of between 3-4 years [7, 8]. Moreover, COVID-19 may have reduced numbers of cases because of reduced travel or by decreased reporting.

Characteristics of cholera’s epidemiological patterns have been described [9–12]. The Spatio-temporal characteristics of the 2010 outbreak that started in Borno State and grew in magnitude to spread to entire Northern Nigeria in three waves appeared to be amplified by flooding [9]. Modeling of hospital case data from Kano, Sokoto, and the Zamfara States between 1991 and 2011 demonstrate two peaks of disease transmission between April and August [9–12]. Increases in temperature, rainfall, poverty, and population density were found to be associated with both cholera cases and deaths [9].

Despite the rich literature on cholera outbreaks in Nigeria and Kano State specifically [13, 14], there is limited understanding of cholera hotspots, the geographically limited areas where socio-cultural and environmental conditions facilitate the transmission of the disease, and where cholera persists. In 2017, the Global Task Force on Cholera Control (GTFCC) called for the elimination of cholera in 20 countries by 2030 [15]. To achieve this goal, GTFCC emphasized detecting and directing interventions including oral cholera vaccine (OCV) to cholera hotspots. In 2018, Nigeria established the National Strategic Plan of Action on Cholera Control (NSPACC) to pre-emptively vaccinate populations in identified hotspots nationwide between 2018 and 2023 [16]. The current hotspot maps in the NSPACC, as well as those recently published for Kano State [8], are all based on the Local Government Areas (LGA). However, the LGA is a large area, and hotspot maps at this level may be too large to be feasible. Specifically, cholera risk does not appear to be uniform within an LGA; rather, higher risk areas likely exist in smaller pockets within the LGAs. We hypothesize that cholera hotspot maps at the ward level would be more helpful in targeting interventions including OCV, but for which we found no studies delineating cholera hotspots at the ward level (hotspot wards). In this study, hotspot wards are termed micro-hotspots. The ward is the fourth and smallest administrative level in Nigeria; thus, studying cholera at this level will provide an improved understanding of micro-hotspots, the specific wards prone to initiating, sustaining, and spreading yearly epidemics. This is urgently needed to provide decision-making vis-a-vis critical interventions including OCV for cholera elimination by 2030.

To illustrate the difference between an intervention at the level of an LGA compared to a ward, the average population of an LGA of Kano State is about 300,000 [17], and there are 10 to 15 wards per LGA. Administratively, Kano State is divided into 44 LGAs; In turn, the 44 LGAs are further divided into 484 wards (Fig 1). Of these, 88 are urban wards. Though we identified hotspots LGAs in Kano State [8], the specific ward where the interventions need to focus was not identified; in fact, very little is known about cholera transmission at the ward scale. Studies by George *et al*. [18] and by Ali *et al*. [19, 20] illustrate the focal nature of cholera transmission and show that cholera risk is much higher close to index cases than it is at a distance. Attempts to define hotspots at the LGA level [8] miss the true facilitators of transmission that can be identified by studies of micro-hotspots.

**Fig 1.**
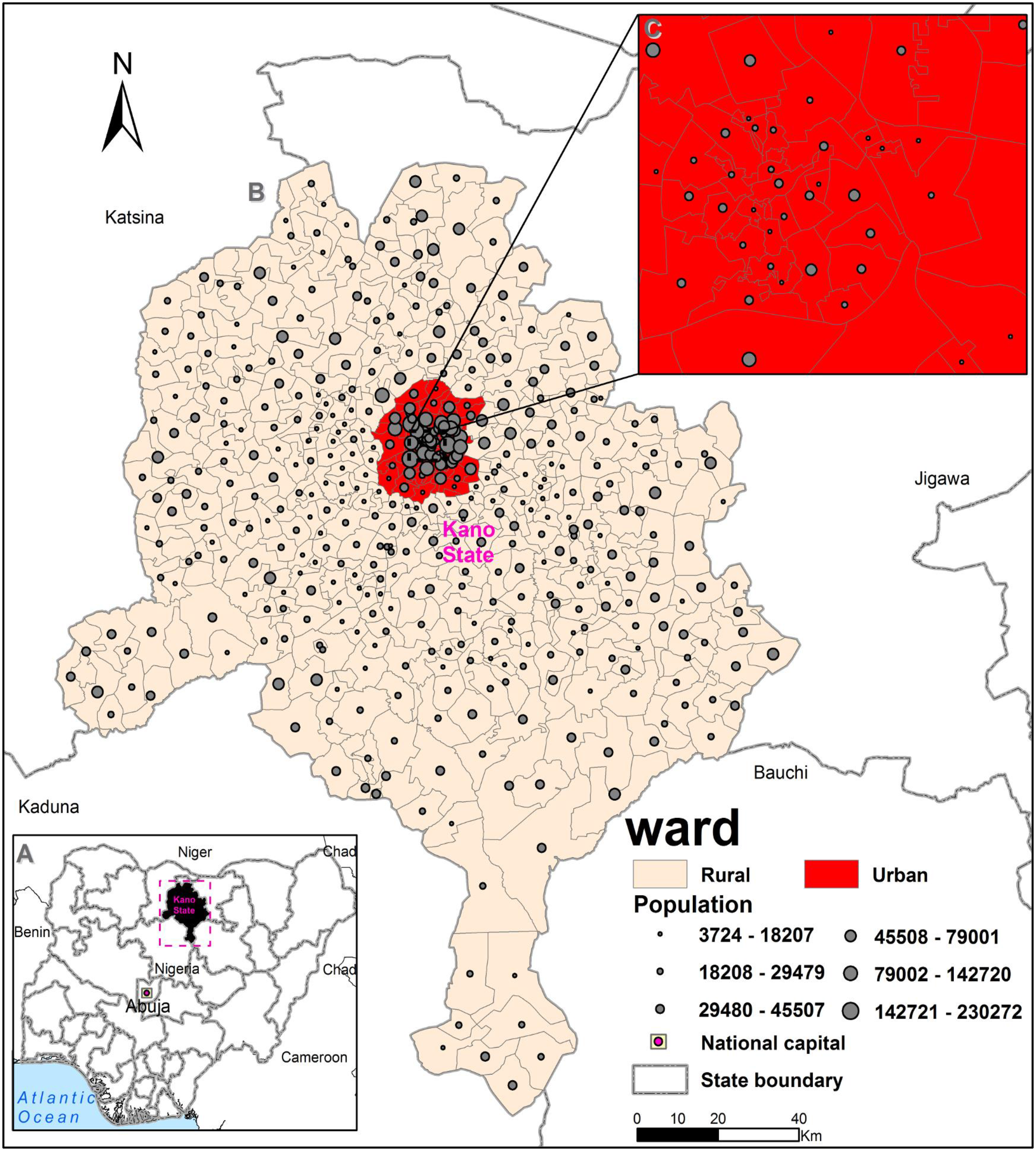
Study setting depicting population distribution in rural and urban wards of Kano State. (A) Insert shows Nigeria with national capital Abuja and Kano State in Northern Nigeria while (B) describes the population distribution of the 484 wards by rural (396 wards) and urban (88 wards) locations. (C) Insert show urban wards at the Centre of the State.

Conceptually, the study of cholera micro-hotspots in Kano State is the “link” between the household level studies of George *et al*. [18] and the district level hotspot mapping of M’bangombe *et al*. [21], Bwire *et al*. [22], and Ngwa *et al*. [7, 8]. The national plan for Uganda included hotspot mapping to identify the districts to be targeted for OCV and water sanitation hygiene (WASH) [22]. However, within the districts, local officials provided additional local, but still subjective, information as to the sub-districts, which were the “true hotspots,” and these sub-districts were then targeted. This study replaces the subjective impressions of local officials with objective information related to rates and risks of cholera at ward level.

Several methods have been described for analyzing the spatial and temporal patterns of disease outbreaks. Ripley’s K function evaluates spatiotemporal patterns of disease occurrence to detect the magnitude of clustering at different temporal and spatial distances [23]. The spatiotemporal Knox test detects statistically significant clusters of disease cases at specific temporal and geographic distances [24], and the Mantel index improves the later method by allocating higher weights to nearby incidents both temporally and geographically [25]. Further, the kernel density estimation (KDE) method describes and visualizes patterns of space-time disease clusters using heat maps [26] while the space-time kernel density estimation (SPKDE) extends the KDE capabilities and visualizes space-time disease clusters using heat volume maps [27]. In this study, we used scan statistics (SaTScan) to analyze and map space-time patterns of cholera clusters/hotspots [28] and also used a method recently described by the GTFCC which utilizes the mean annual rate in a defined area in combination with the persistence of cholera in the area [29].

This study fills a gap for Kano State by describing 1) patient characteristics (suspected and confirmed cases and deaths) including CFR, severity (hospitalization), demographic variables (gender and age), care-seeking behavior; 2) describing laboratory confirmation (stool sample collection and rapid diagnostic tests); while analyzing 3) temporal, and 4) geographical patterns of spread incorporating attack/incidence rates, and 5) describing cholera micro-hotspots. The study aims to compare all five points between the urban and rural settings to identify critically, yet poorly appreciated micro-hotspots of cholera in Kano State, and to do so while employing statistical methods in addition to the tool recommended by GTFCC with health facility-based cholera surveillance data set using data between 2010-2019.

## Methods

### Ethics consideration

We used an anonymized historical line-list dataset at the ward level from the Nigeria Centre for Disease Control (NCDC). The Health Research Ethics Committee of the Ministry of Health, Kano State provided ethical approval of the study (Ref: MOH/Off/797/T.1/2012) as part of an ongoing effort to inform a national plan for disease control and elimination from the State. Based on these, the Johns Hopkins University Internal Review Board (IRB) determined that the study is not human subjects’ research and was exempt from IRB review. Based on the anonymity of data, we did not obtain informed consent.

### Study setting

Nigeria, a Federation of 36 states and the Federal Capital Territory, Abuja, is located along the West Coast of Africa (Fig 1A). The landmass covers the mangrove swamps and creeks in the south that transitions through the tropical forest, savannah woodlands, and grasslands in the middle to the Sahel in the North. Kano State (Fig 1A), our study site, is in the Sahel in the Northwest of the country. With an estimated population of 13,076,900 people in 2016, it is the most populated state of Nigeria [30]. The dominant spoken language is Hausa while English is the official language. In 2019, the average population of an LGA was 325,256 with a range of 149,057 in the rural LGA of Tofa to 909,993 in the urban LGA of Nassarawa. The average population of a ward was 29,547 ranging from 3,724 in the rural ward of Kwami (Tofa LGA) to 230,272 in the urban ward of Dorayi (Gwale LGA) (Fig 1B). The population is highest in the Center urban wards (Fig 1 C) and progressively decreases outwards towards the rural wards. Among the states of Nigeria, we chose Kano State because of the interest of NCDC and Kano Ministry of Health in cholera control and because of yearly cholera outbreaks [13, 14, 31]. In addition, Kano State has scheduled an OCV immunization program to hold between 2020 and 2023; and thus, this analysis will provide data to inform decision-making on the highest priority micro-hotspots.

### Data collection

We obtained a de-identified cholera line-list (Microsoft Excel, 2010) of both confirmed and suspected cases at ward level for Kano State from NCDC spanning the period from 2010-2019. Data from the line-list included age, gender, date of symptom onset, date seen at the health facility, whether inpatient or outpatient, the outcome of visit (survived or died), stool sample collection and testing, as well as the ward of patient’s residence. The cholera case definition [32] used for the line-list was based on the Integrated Disease Surveillance and Response strategy, the public health surveillance system used to capture data at health facilities across Nigeria [32, 33]. During visits to a health facility, data on cholera cases are captured and entered into the line-list. The data is de-identified and reported to the ward and LGA Disease Surveillance and Notification Officers where data are compiled and sent to the Epidemiology Unit of the State Ministry of Health. The state analyzes the data and sends these to the NCDC, the national public health institute that coordinates surveillance activities, and from there to other government entities and international partners.

The last population and housing census for Nigeria were conducted in 2006 [35] but did not provide population sizes at the ward level. Hence, we obtained population sizes of each ward between 2010 and 2019 from the Expanded Programme on Immunization (EPI) in Nigeria [34, 35]. EPI’s population estimates are considered accurate as the program conducts headcounts of the population per ward to get accurate population denominators for immunization. The EPI estimates that 49% of the Kano State population is female, so we used this proportion to estimate female populations in urban and rural locations. Spatial data such as polygon Shapefiles at the state, LGA, and ward levels were obtained from public health authorities in Nigeria, and classification of wards as urban or rural was read directly from the ward Shapefile. Furthermore, (inland water bodies) data on river lines and lakes, i.e., the contextual factors of cholera transmission, were obtained online (http://www.diva-gis.org/Data). All spatial data in the geographic coordinate system were projected into the Universal Transverse Mercator, Zone 32N, coordinate system.

### Data cleaning/wrangling

The line-list cholera data set for each year from 2010-2019 contained several columns including the ward name of the patient. As this study focused on analyzing cholera at the ward level, the ward name was critical in capturing and mapping the ward of infection. We observed that about half of ward names found in the ward column were not the official administrative ward names found in the Shapefile, which does provide the official ward names. They were rather names of villages/settlements/schools or tribes/ethnic groups or local chiefs/community leaders or even roads/streets/buildings. To obtain the official administrative ward names, researchers at Johns Hopkins University worked with the Kano State Epidemiologist, who, in turn, worked with the LGA and ward representatives to infer the official ward names from the ones in the line-list. For example, one of the unofficial ward names in the 2011 line-list was Civic Centre, which is the name of a building and as such not an official ward name. So, working with the State Epidemiologist, LGA, and ward representatives, the unofficial ward name Civic Centre was replaced with Fagge A, as the Civic Centre building is located in Fagge A ward. We also used an online resource: (https://en.wikipedia.org/wiki/List_of_villages_in_Kano_State) to infer some of the official ward names based on the names of villages and schools mentioned in the line list. Unfortunately, we were unable to trace all the unofficial ward names to official ward names because they were either misspelled, abbreviated or blank; in which case, they were classified as missing. To illustrate, for the 2011 line-list, out of 785 records, 456 (58.1%) ward names were unofficial but were traced and replaced with official ones while 17 (2.2%) could not be traced, and as such were classified as missing. This process was repeated with the line-list data for each year from 2010-2019.

Dates of disease onset and health facility visits in the ward line-list were in various formats from one year to the next. These were all standardized to the year-month-day format. Also, the ward level Shapefile had missing polygons; so, to add the missing polygons, we created new Shapefile polygons with the Geographic Coordinate System and digitized the ward polygon boundaries in ArcGIS Version 10.5.1 (ESRI, Redlands, CA, USA). Finally, some wards in the Shapefile that belonged to the rural location were misclassified as urban and vice-versa and this was corrected with input from the State Epidemiologist.

### Data analysis

#### Descriptive temporal, geographical, and person variation analysis

We used propagated epidemiological curves to depict weekly trends of cholera cases and CFRs (%) in our study time period from 2010 to 2019 (S1 Fig A and S1 Fig B). To visualize the geographic distribution of the ward level cholera crude AR per 100,000 population, the underlying risk of cholera infection within a ward, choropleth maps were produced for the entire study period. Crude ARs were computed for each ward using the formula number of reported cholera cases in a ward during a year divided by annual population estimate for 2010 to 2019 in R version 3.6.2 [36]. To address concerns that patterns in the yearly ward crude ARs may be unstable (i.e., variance instability) [37–40], Empirical Bayes Smoothed (EBS) rates were estimated in GeoDa Version 1.16.0.12 [41], and compared to the corresponding crude ARs via box plots (S2 Fig) and visualized using choropleth maps. Generally, variance instability is due to 1) small number of reported cases or 2) small areas or 3) small population sizes [37], all of which were applicable with our ward data. However, because of minor differences between the crude and EBS attack rates (S2 Fig), marginal differences in the results using EBS rates and crude ARs (S3 Fig), and EBS rates showing cholera in wards not known to report the disease, we proceeded with using the crude ARs.

To describe the dynamics of yearly cholera outbreaks for 2010-2019, we stratified the cases and deaths by location (urban/rural), gender (female/male), and age groups (<5, 5-14, >15 years). We then calculated the overall attack (suspected and confirmed cases/population) and death (deaths/population) rates, CFRs (deaths/suspected and confirmed cases), proportion of suspected cases and deaths, proportion of cases classified as severe (hospitalized cases), proportion of female, proportion of stool samples taken and positive rapid diagnostic tests, and proportion of rapid care-seeking. Severity was defined as hospitalization due to or death from cholera. A table and histogram were produced to visualize the distribution of cholera suspected cases and deaths as a function of location, and age group. We used the Chi-square test to analyze associations between location, gender, and age group, and outcome (survived/died) of cholera. *P-values* of the test statistic were reported with 5% (i.e., α = 0.05) significance level.

#### Laboratory confirmation analysis

Laboratory surveillance collected stool samples from suspected cholera patients when they presented to health facilities with symptoms of cholera to test the presence of *V. cholerae* using rapid diagnostic tests (RDTs) (Crystal VC, Span Diagnostic, India). To confirm the results of clinical diagnosis and RDTs, samples with positive RDTs were forwarded to the NCDC reference lab in Abuja for analysis using standard culture techniques [42]. Information on samples taken and RDT tests were read from the ward line-list for 2010 to 2019; however, culture results were not available. We presented the results in a table.

#### Space-time cluster detection analysis

In this study, we used scan statistics (SaTScan) to analyze and map space-time patterns of cholera clusters/hotspots across Kano State at the ward level incorporating weekly disease incidence for 2010-2019 [28]. This is a common method, which ascertains whether the number of reported cases of the disease in a ward exceeds the expected number compared to rates outside the ward. We also used a method recently developed by the GTFCC but applied it to the geographic area of the ward rather than the LGA (district) [29]. We chose SaTScan primarily because it was commonly used in detecting clusters of disease incidence including cholera hotspots detection [7, 22, 43, 44]. Secondly, SaTScan incorporates spatial and Spatio-temporal scan statistics with linkage to GIS for results visualization. We also used the GTFCC tool [29] to assess how it compares with SaTScan in hotspot identification and classification. The GTFCC tool, based on Microsoft Excel, can readily be used with ease in resource-poor settings at the periphery level (ward and health facility level) by health personnel without statistical/GIS skill competencies needed to use SaTScan and GIS.

#### Cholera clusters/micro-hotspots detection analysis based on SaTScan method

Clusters and micro-hotspots of cholera reported cases were detected using SaTScan v.9.6 through a retrospective analysis of the data for 2010-2019. To assess the dynamics of cluster patterns in this time period including variations in size and duration as well as how clusters of outbreaks moved through space and time, a Poisson discrete space-time scan statistic was implemented following Kulldorf *et al*. [28]. We scanned for high rates with 25% spatial and temporal windows, respectively, and default settings for all other values. The data used in this analysis were weekly reported cholera cases for the ten years and the annual population sizes. The space-time cluster dynamics were presented in a table and visualized using ArcGIS Version 10.5.1 (ESRI, Redlands, CA, USA). The most likely cluster (micro-hotspots) is the ward that is least likely to have occurred by chance. In keeping with our objective of comparing micro-hotspots detection using SaTScan and the newly developed GTFCC tool, the relative risks greater than 1 of the Spatio-temporal scan statistic with a p-value of 5% or less were shown in a table and mapped using ArcGIS Version 10.5.1 (ESRI, Redlands, CA, USA).

#### Cholera micro-hotspot detection analysis based on the GTFCC method

Applying the GTFCC tool [45], we identified and ranked cholera micro-hotspots based on two epidemiological indicators 1) mean annual incidence per 100,000 population of reported suspected and confirmed cases, and 2) cholera persistence in the entire study period. The indicator mean annual incidence was computed by first calculating the annual incidence by dividing the number of reported cholera cases of each ward by its population size for each year. Next, we calculated the mean of the annual incidences for the ten-year study period for each ward. For persistence, we divided the total number of weeks with reported cholera cases by the total number of weeks in the study period. All calculations were performed in Microsoft Excel 2016 [29].

Types of micro-hotspot (T) were assigned to each ward based on the two indicators delineating priority levels into high (T1), medium (T2), medium (T3), and low (T4). A 55^th^ and 71.6^th^ percentile values were defined as the cut-off points for high mean incidence rate and persistence; however, the cut-off levels could be adjusted as desired. T1 micro-hotspots corresponded to high priority areas and were wards with high mean annual incidence and high persistence of cholera during the surveillance period. T2 micro-hotspots corresponded to medium priority areas, which were wards with a high mean annual incidence rate and low persistence of cholera. T3 micro-hotspots corresponded also to medium priority areas characterized by a low mean annual incidence rate, but a high persistence of cholera. T4 micro-hotspots corresponded to low priority wards, which had a low mean annual incidence rate and low persistence of cholera. The cut-off values for incidence and persistence delineating priorities/types of micro-hotspot were chosen based upon the objectives of the NSPACC for Nigeria for 2018–2023 and resources available for effective implementation of intervention measures. Cholera hotspots classification chart and map were produced using the R statistical computing environment (version 3.6.2) [36] and ArcGIS, respectively.

## Results

### Descriptive person, temporal, and geographical variations

#### Descriptive person (patient) characteristics variation

In our study period, 18483 suspected and confirmed cholera cases and 617 deaths (CFR = 3.3%) were reported in urban and rural settings from Kano State. Of these, 67.7% (12580/18483) of the cases for the study period were from rural wards; however, overall AR was slightly higher in the urban wards. Still, 72% (443/617) of the deaths were from rural wards, which saw a slightly higher death rate (Table 1). CFR decreased with increasing age ranging from 7.4% among children under five years old to 2.6% among the working-age group (≥15 years) in the urban area. Cholera severity (hospitalization or death) was higher in the urban (77.4%) compared with the rural (53.4%) setting and the highest severity (84.7%) was registered among people 15 years and older. In addition, there were more females with cholera overall (51.5%) than males from 2010 to 2019, especially in the rural setting among women 15 years and over (59.3%). Stool samples tested using RDTs were obtained from only 1.3% of all cases in this study period and were least often obtained from children 5-14 years old (0.7%) in the rural area and were more often obtained from those under 5-year (3.5%) in the urban location. Of the stool samples collected, 62.7% were positive for *V. cholerae* using RDT. The positivity rate was lowest in the urban wards amongst children under age 5 years (41.2%), but highest in the rural wards amongst children of the same age group (68.4%). Finally, nearly 75% of all reported cases from 2010-2019 sought clinical care on the same day of symptom onset with a range of 52.1% in the urban to 82.7% in the rural setting (Table 1).

**Table 1.**
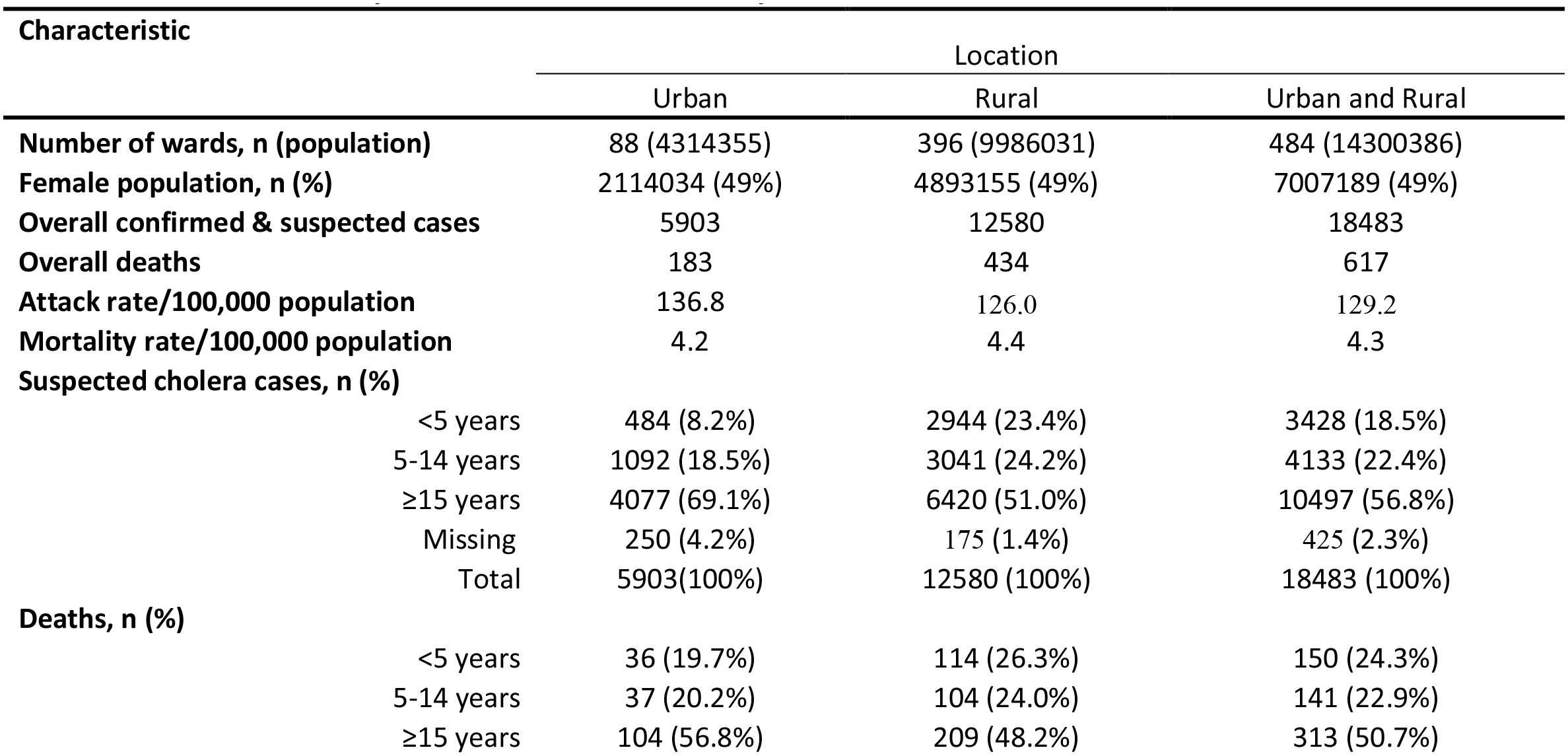
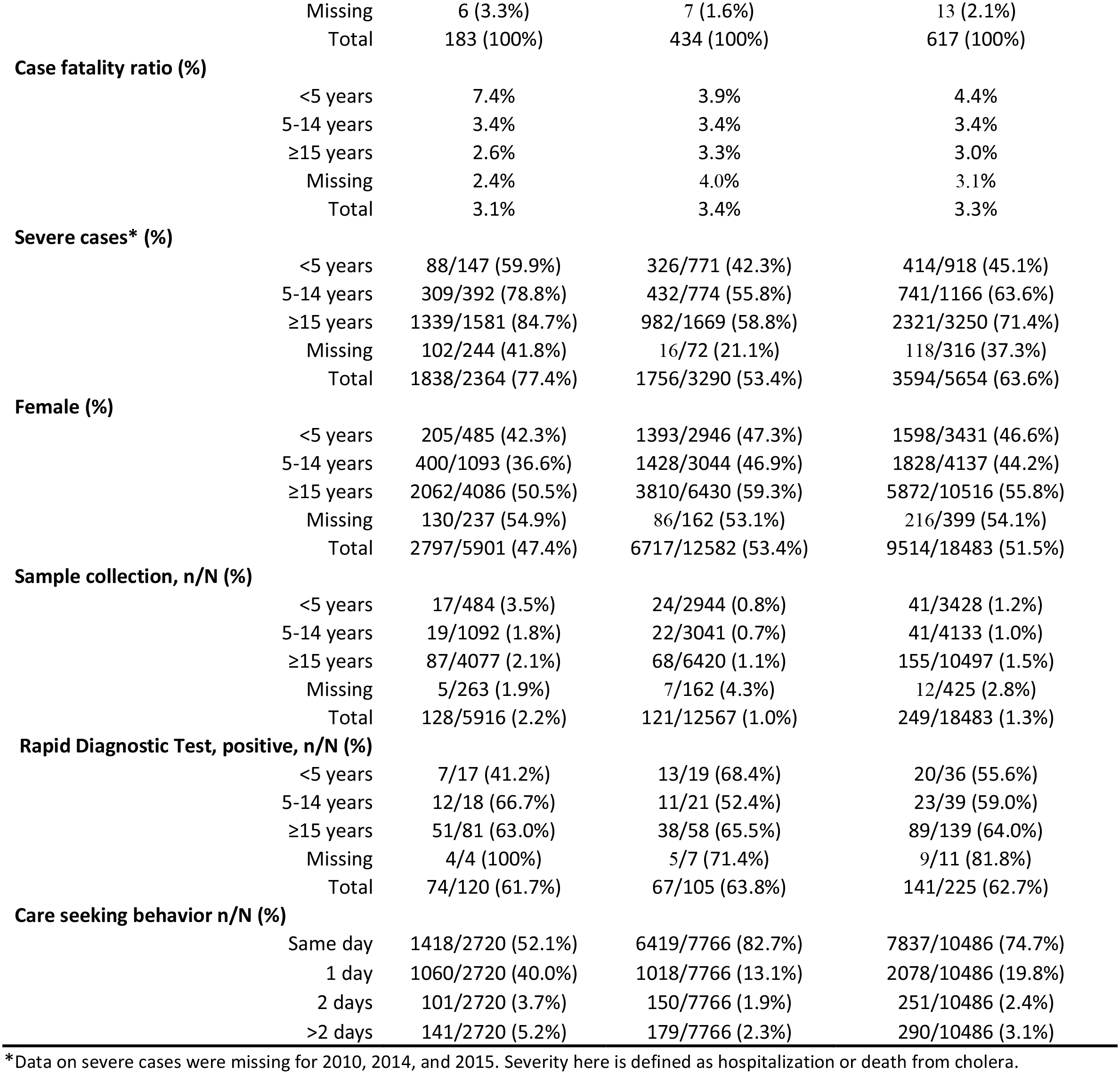
Characteristics of suspected and confirmed cholera patients in Kano State, 2010-2019.

From 2010-2019, the ages of the cases ranged from 1 month to 100 years. In this period overall, we found statistically significant evidence that cholera outcome (survived/died) were gender (*p-value* = 0.0006), and age group (<1-4, 5-14, and >15 years) (*p-value* = 0.0005) dependent. Although we found that location was not associated with overall outcome (*p-value* = 0.2465), yet, amongst women, cholera outcome was statistically and significantly associated with location (*p-value* = 0.003). In contrast, amongst men, cholera outcome was **<u>not</u>** linked with location (*p-value* = 0.4527). Excluding 2011 and 2013, there were more cases and deaths among women in the rural setting than those in the urban setting (Fig 2). Apart from 2010 and 2011 in the urban setting, the annual age distribution of suspected and confirmed cholera cases and deaths showed a right skew shape (Poisson distribution); i.e., there were more cases of cholera among people in younger age brackets (<5 and 5-10 years old) compared to any other age bracket (Fig 2).

**Fig 2.**
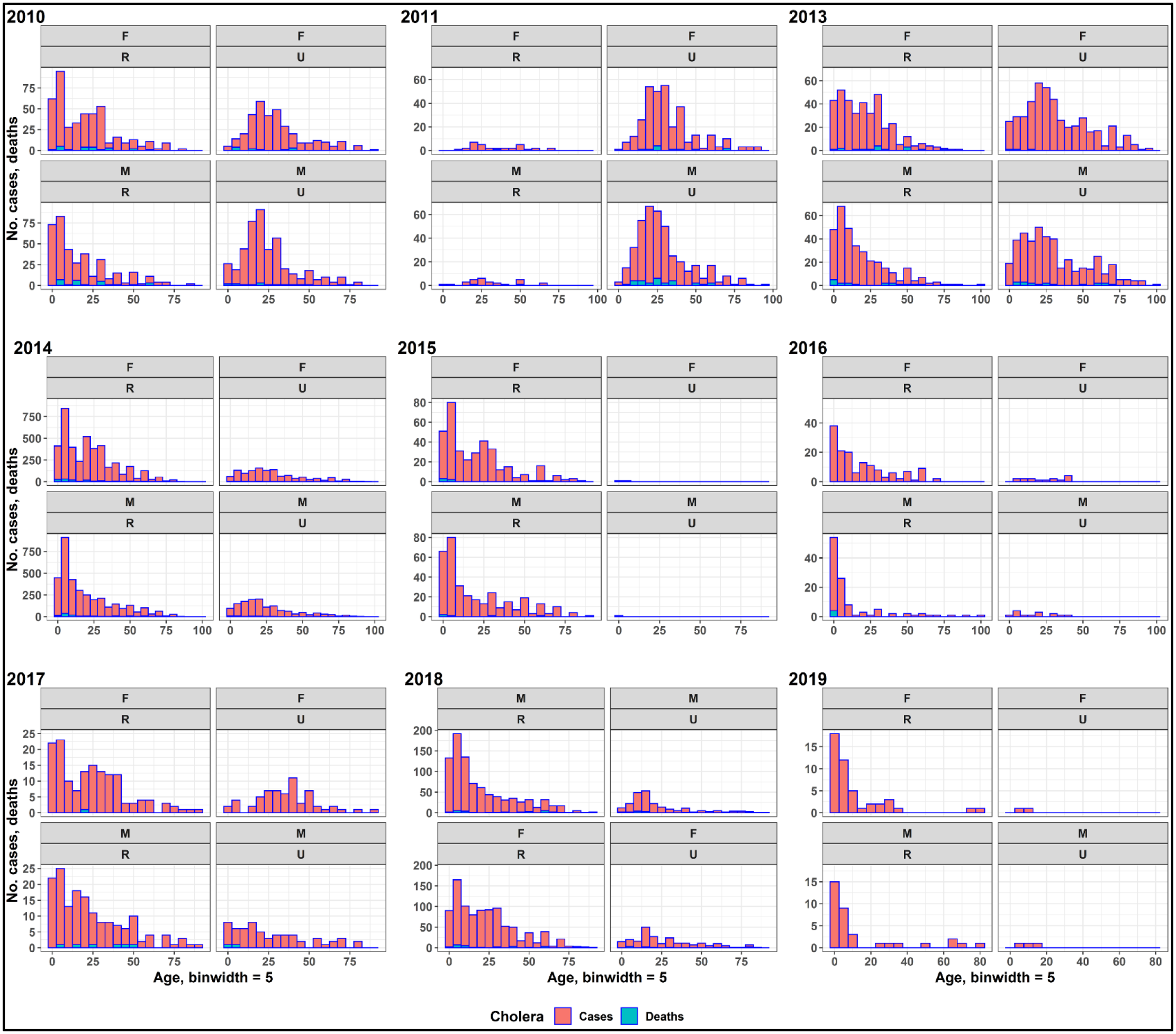
Suspected and confirmed cases and deaths distribution of cholera by age, gender, and location, 2010-2019. <u>Of note</u>, binwidth = 5 means cases and deaths distribution based on age groups of five years interval. 2010 to 2019 represents years from 2010-2019. (F) Female, (M) Male, (R) Rural, and (U) Urban are the stratifications of gender and location of infection, respectively. This figure depicts the urban and rural divide of cholera outbreaks annually. Apart from the years 2010 and 2011 in urban setting, the distribution of cholera cases by age has a right skew (Poisson) shape. This implies that, irrespective of sex and location, the cases of cholera were higher among people of younger ages (<10 years) than older ones. Excepting the years 2011 and 2013, amongst women, cholera survival significantly depended on setting (*p-value* = 0.002); Women in rural areas were more likely to die from cholera than women in urban areas. These findings confirm that age, gender and location are associated with cholera outcomes (survived or died).

#### Descriptive Spatio-temporal variation of cholera

Weekly reported cholera cases in our study period started in the rural setting during week 31 of 2010 during the rainy season (Fig 3, S1 Fig A). Weekly CFRs ranged from 80% during week 31 of 2010 to 9% during weeks 17 and 31 of 2015, although there were no cases reported in 2012 and no fatalities in 2019 (S1 Fig B). In 2014, over 450 cases were reported in the rural setting among women during week 27 alone (as we have seen above, generally, weekly cases were greater among women in rural locations). Excepting the year 2013, cases in the rural and urban settings peaked asynchronously (Fig 3), i.e., when cases in the rural setting were at their peaks, those in the urban location were at their troughs. Further, cholera cases occurred in both the rainy and dry seasons. For instance, in 2011, 2013, and 2019, cases occurred predominantly in the dry season at either the start or end of the year. However, during the other six years studied, large numbers of cases occurred during the rainy season from week 22 (early June) to week 40 (late September) with peaks in August (Fig 3). The spatial distribution of wards with high cholera crude AR per 100,000 population revealed considerable spatial heterogeneity between 2010 and 2019 with cholera occurring in the central, north, south, east, and west portions (Fig 4 CR). We found marginal differences between spatial distribution in the cholera crude ARs and EBS rates (Fig 4 CR, Fig 4 EBS). The widest spatial spread occurred during 2014 and 2018, but in no year were all wards affected even after smoothing (Fig 4).

**Fig 3.**
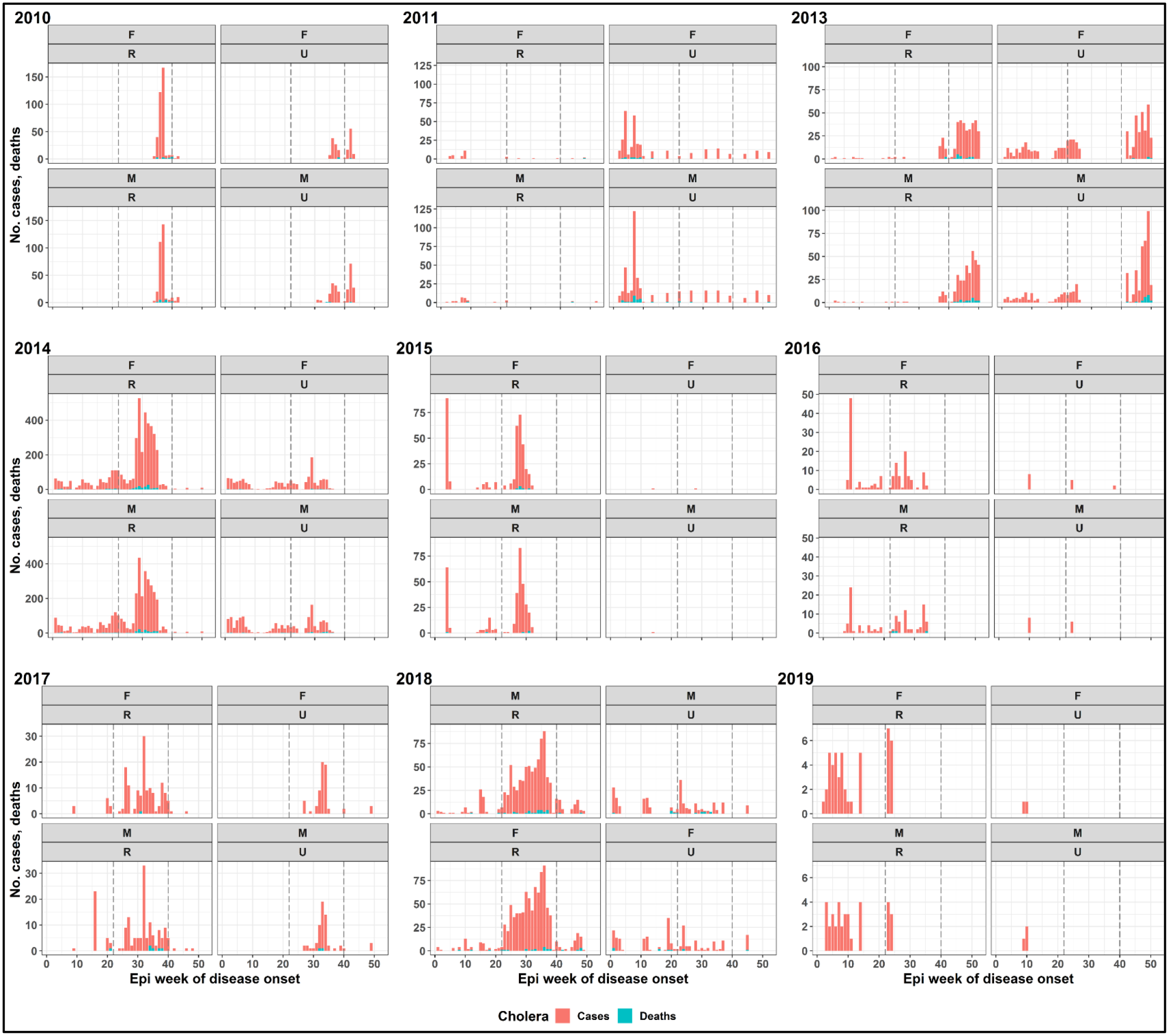
Weekly occurrence of cholera epidemics in Kano State, Nigeria, 2010-2019. The y-axis represents suspected and confirmed reported cases and deaths of cholera while the x-axis shows the epidemiological (Epi) week of disease onset. F (female), M (male), R (rural), and U (urban) represents the gender and location stratifications. The dashed gray lines indicate the rainy season period (from June to September). In our analysis time period, the outbreaks started among men in the rural setting in week 31 of 2010 during the rainy season. Generally, weekly transmission was greater among women in rural location. Excepting the year 2013, outbreaks in the rural and urban settings peaked asynchronously, i.e., when outbreaks in the rural setting were at their peaks, those in the urban location were at their troughs. Further, cholera exacerbations occurred in both the rainy and dry seasons. For instance, in 2011, 2013 and 2019, epidemics occurred predominantly in the dry season at either the start or end of the year. In the rest of the six years studied, intense transmission of cholera occurred during the rainy season from week 22 (early June) to week 40 (late September) with peaks in August.

**Fig 4.**
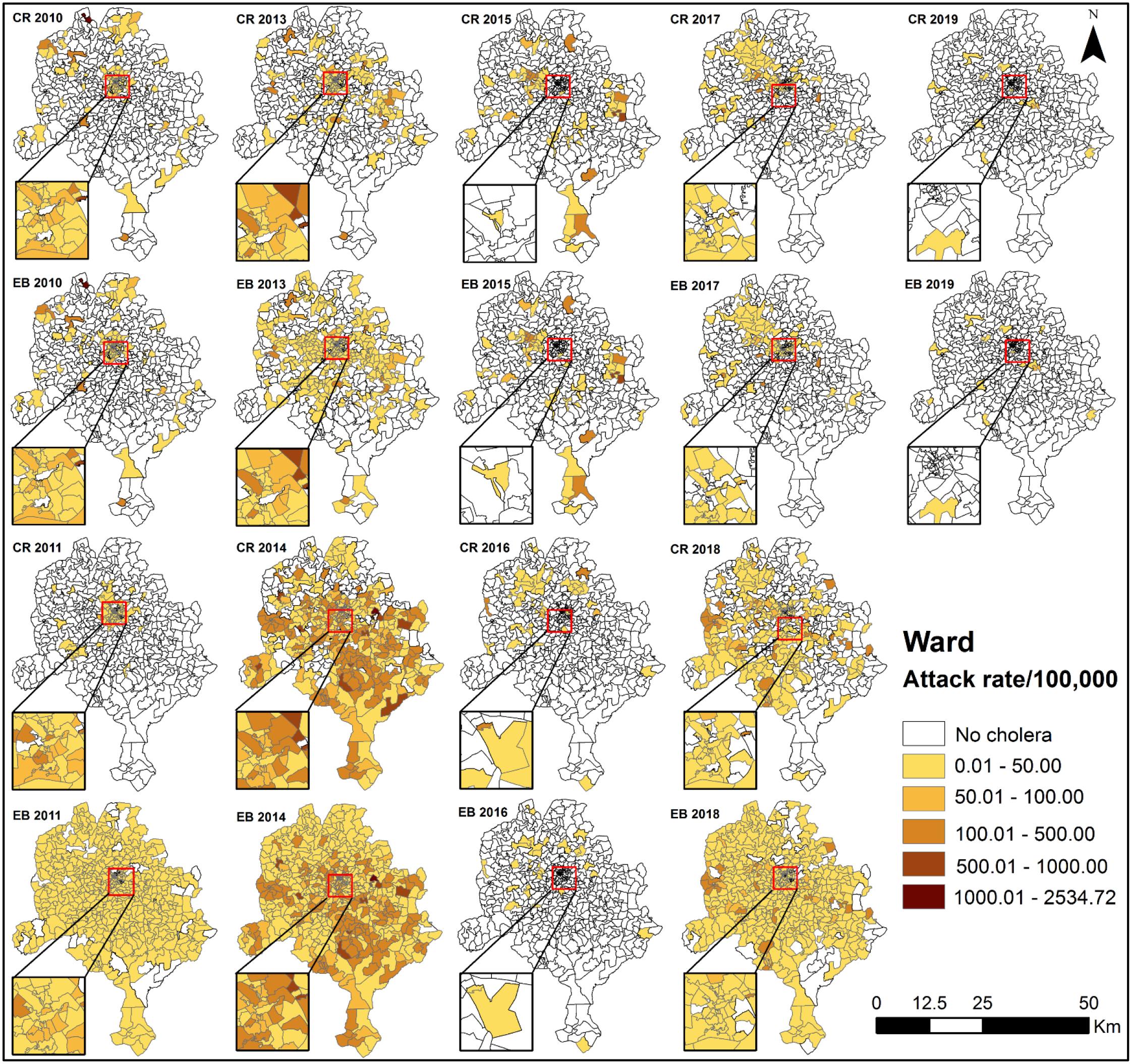
Distribution of ward crude and Empirical Bayes Smoothed cholera attack rates from 2010-2019. The map shows differences between the distribution of crude rates (CR) and Empirical Bayes Smoothed (EBS) rates. The largest difference between CR and EBS occurred in 2011. Nevertheless, because the EBS rates show cholera occurring in wards where cholera is not known to occur or where the disease has not been reported, we elected to use the CR in the rest of the analysis. There were no data for 2012.

### Cholera clusters and micro-hotspot detection

#### SaTScan Spatio-temporal cluster and micro-hotspot detection

The results of SaTScan cluster detection at the temporal scale of week and spatial scale of ward between 2010 and 2019 yielded three statistically significant cholera clusters of reported cases involving 254 wards (Table 2), as visualized in Fig 5. The pattern of spread of the three clusters shows extensive variation in space and time within Kano State. One cluster occurred between week 36 and 37 of 2010 with a center in the ward of (3) Kunchi in the Northern portion of the State and another between week 45/2013 and week 35/2014 centered at (2) Karofin Yashi ward at the Center of the State. Between weeks 28-35 of 2014, a very large (1) cluster was reported in the southeast portion of Kano State with a center located in the ward of Bagwaro. Within the three detected clusters, clusters 1 and 2 had the most observed cases involving wards in both urban and rural settings (Fig 5). Cluster 3 has 0 Km radius implying it was contained within the Kunchi ward. Between 2010 and 2019, cluster 3 (located in rural setting) had the shortest duration lasting two weeks while cluster 1 had a longer duration spanning eight weeks. Further, cluster 2, involving both urban and rural settings, had the longest duration and persisted for 43 weeks from 2013 to 2014 (Table 2 and Fig 5).

**Fig 5.**
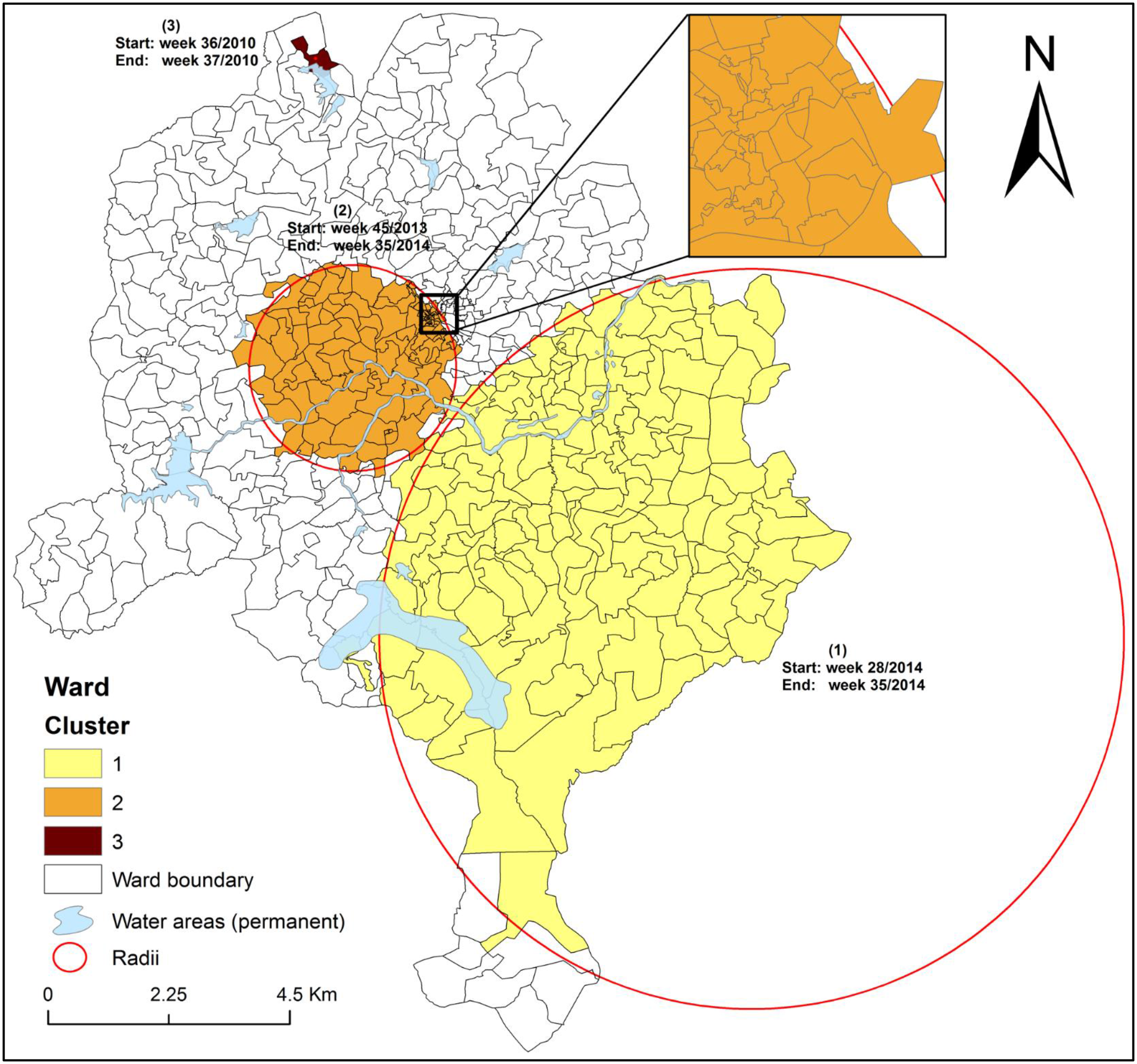
Significant space-time clusters of weekly reported cholera cases in Kano State, 2010-2019. The map depicts dynamics of space-time clusters of cholera at the fine scale of ward, which varied considerably in size and duration. Numbers 1-3 indicate clusters. Clusters (1) and (3) involve rural wards while (2) include urban wards. The big circle shows a (1) cluster with radius of 81.57 Km involving 136 wards while the small circle indicates a (2) cluster of radius 22.70 Km involving 117 wards. Cluster (3) has zero radius, as indicated by the dot (red). Analysis used space-time windows of 25% of the population at risk of cholera and 25% of the study time period, respectively, and default settings elsewhere. Here cholera cluster means a geographically limited area where environmental, cultural and/or socioeconomic conditions facilitate the transmission of the disease and where cholera persists or reappears regularly. The insert shows a better view of the urban area clusters.

**Table 2.** Significant cholera clusters.

| Cluster number* | Duration week/year | Number of wards | P_value | Observed cases | Expected cases | Relative Risk (RR)^ | Population |
| --- | --- | --- | --- | --- | --- | --- | --- |
| 1 | 28-35/2014 | 136 | $1.0 \times 10^{-17}$ | 3098 | 67 | 56.66 | 3069293 |
| 2 | 45/2013 - 35/2014 | 117 | $1.0 \times 10^{-17}$ | 3152 | 355 | 10.61 | 3080827 |
| 3 | 36-37/2010 | 1 | $1.0 \times 10^{-17}$ | 289 | 1 | 4470.32 | 13964 |
| **Total** | — | **254** | — | **6539** | **423** | — | **6,164,083** |

\*Cluster 1 is the most likely while clusters 2 and 3 depict secondary clusters. ^RR is the estimated risk of cholera infection within the cluster divided by the estimated risk outside the cluster.

In Fig 6 and Table 3, we show the relative risk (RR) greater than 1 for each ward associated with significant Spatio-temporal clusters in the period under consideration; and thus, the so-called micro-hotspots, i.e., the specific hotspots wards where cholera occurs more commonly. Of the 254 wards (out of 404) assigned to a cluster (Fig 5), 168 yielded relative risk greater than 1 (greater observed than expected cases) (Fig 6). A breakdown of the 168 micro-hotspots of RR > 1 is in order (Table 3, Fig 6). The (1) largest cluster in the southeastern portion of the State contained 60 micro-hotspots, the larger cluster (2) at the center location spanned 53 micro-hotspots, and the (3) lone cluster in the north involved one micro-hotspot. Fifty-four (54) of the micro-hotspots were not within any cluster. In addition, wards with elevated cholera burden (micro-hotspots) clustered predominantly around contextual factors of cholera transmission namely lakes and rivers (Fig 6). The RR of the 168 statistically significant micro-hotspots ranged from 1.01 in Fanda to 18.63 in Fagge B with a total population of 4,876,254 (Table 3). Furthermore, 79 micro-hotspots with a population of 2,119,974 had a RR ≥ 2 compared to the rest of the State as a whole.

**Fig 6.**
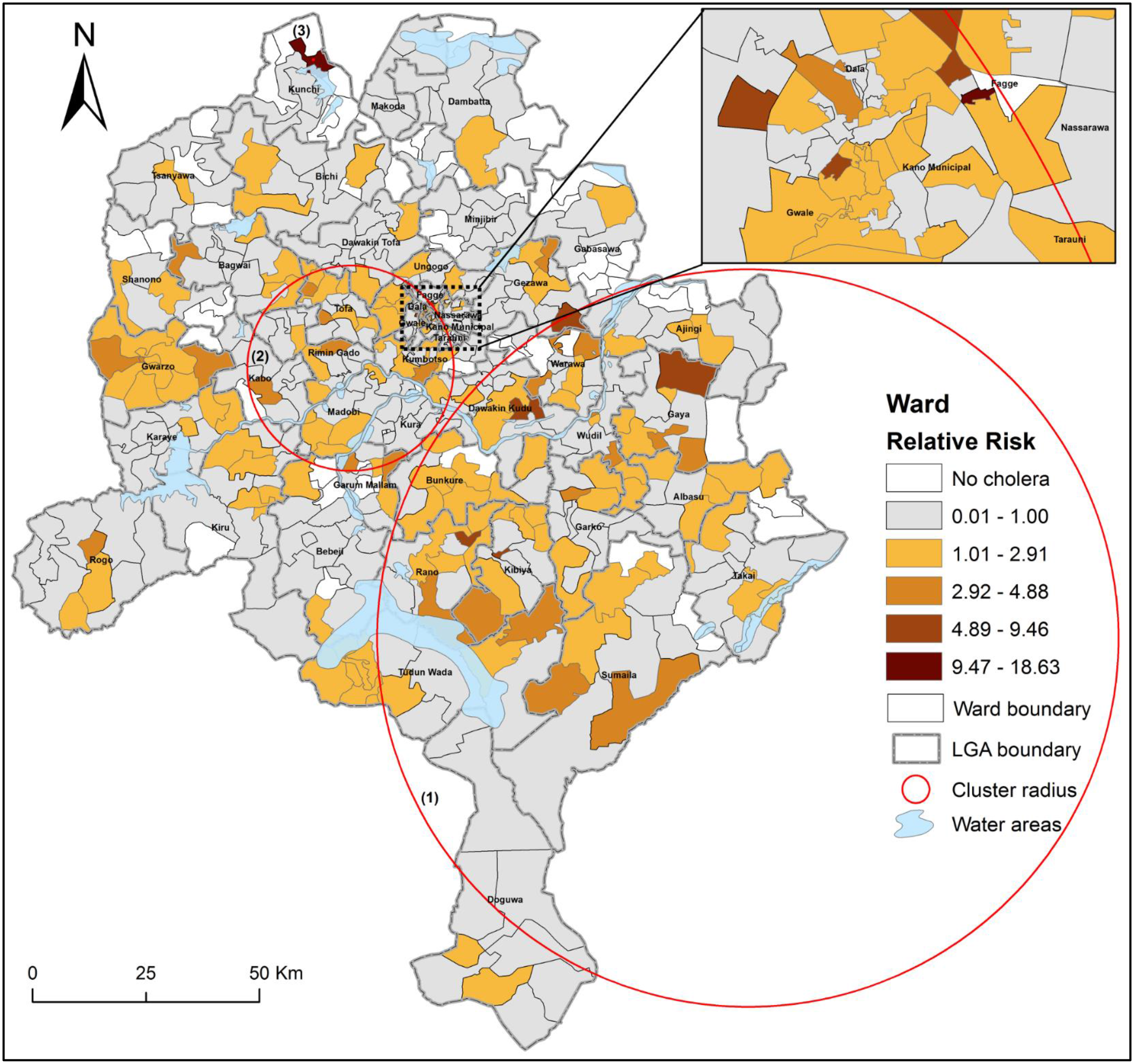
Statistically significant cholera micro-hotspots (ward) in Kano State, 2010-2019. Map depicts relative risk (RR) per ward for the cholera clusters. The micro-hotspots wards (hotspots at the geographical resolution of ward) are those with RR > 1; and thus, wards with statistically significant risk of cholera infection. Of the 448 wards in Kano Sate, we identified ***<u>168</u>*** (37.71%) with RR > 1; and thus, the so-called micro-hotspots. The ward specific RR was calculated within each cluster using space-time scan statistics incorporating reported cholera cases for each week and the ward population for each year. Here cholera micro-hotspot means a geographically limited location where environmental, cultural and/or socioeconomic conditions facilitate the transmission of the disease and where cholera persists or reappears regularly. The insert shows a better view of the micro-hotspots at the Centre location. <u>Note</u> that the labels in the map represents Local Government Areas (LGAs) in which the wards are found.

**Table 3.**
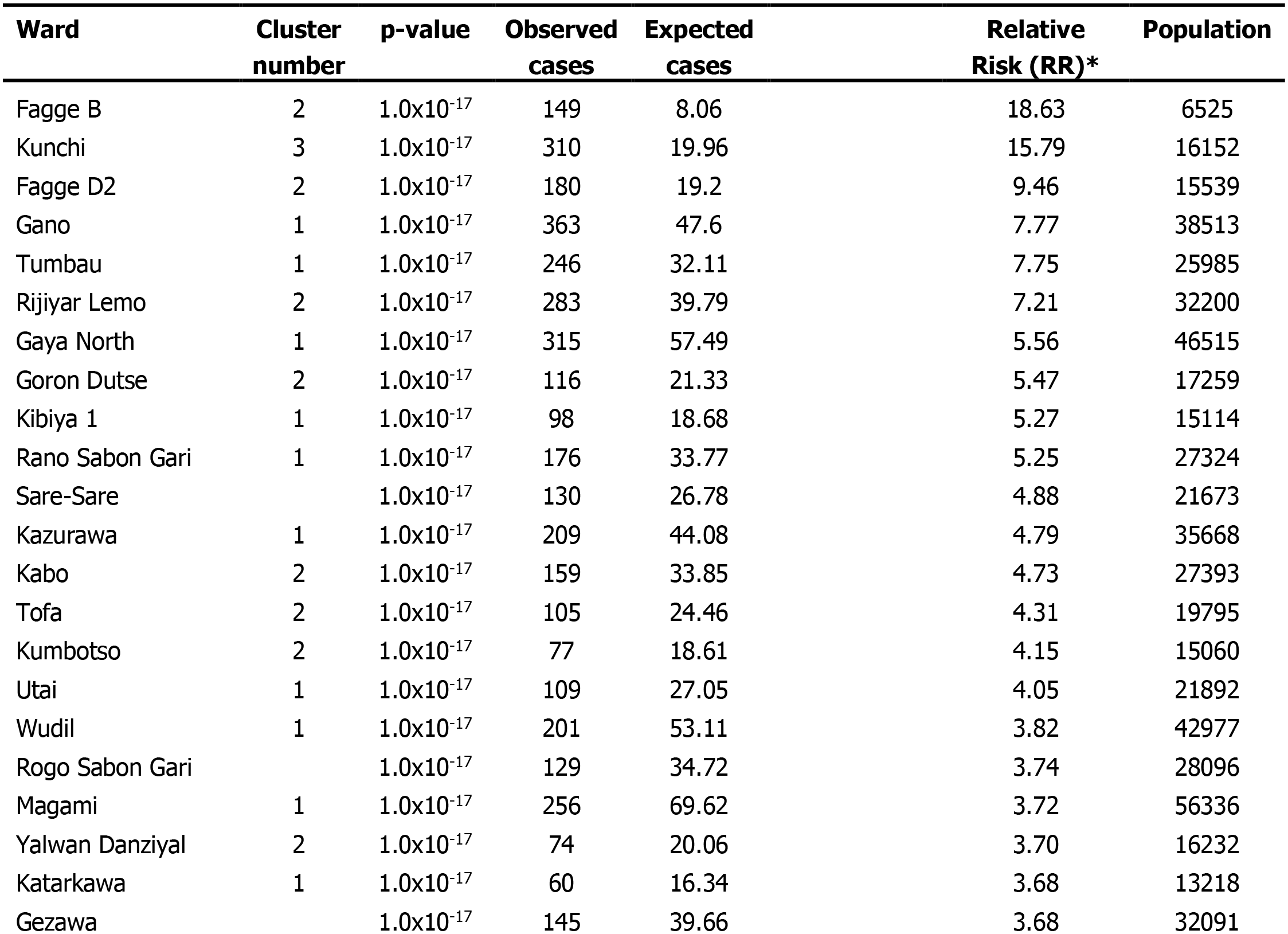
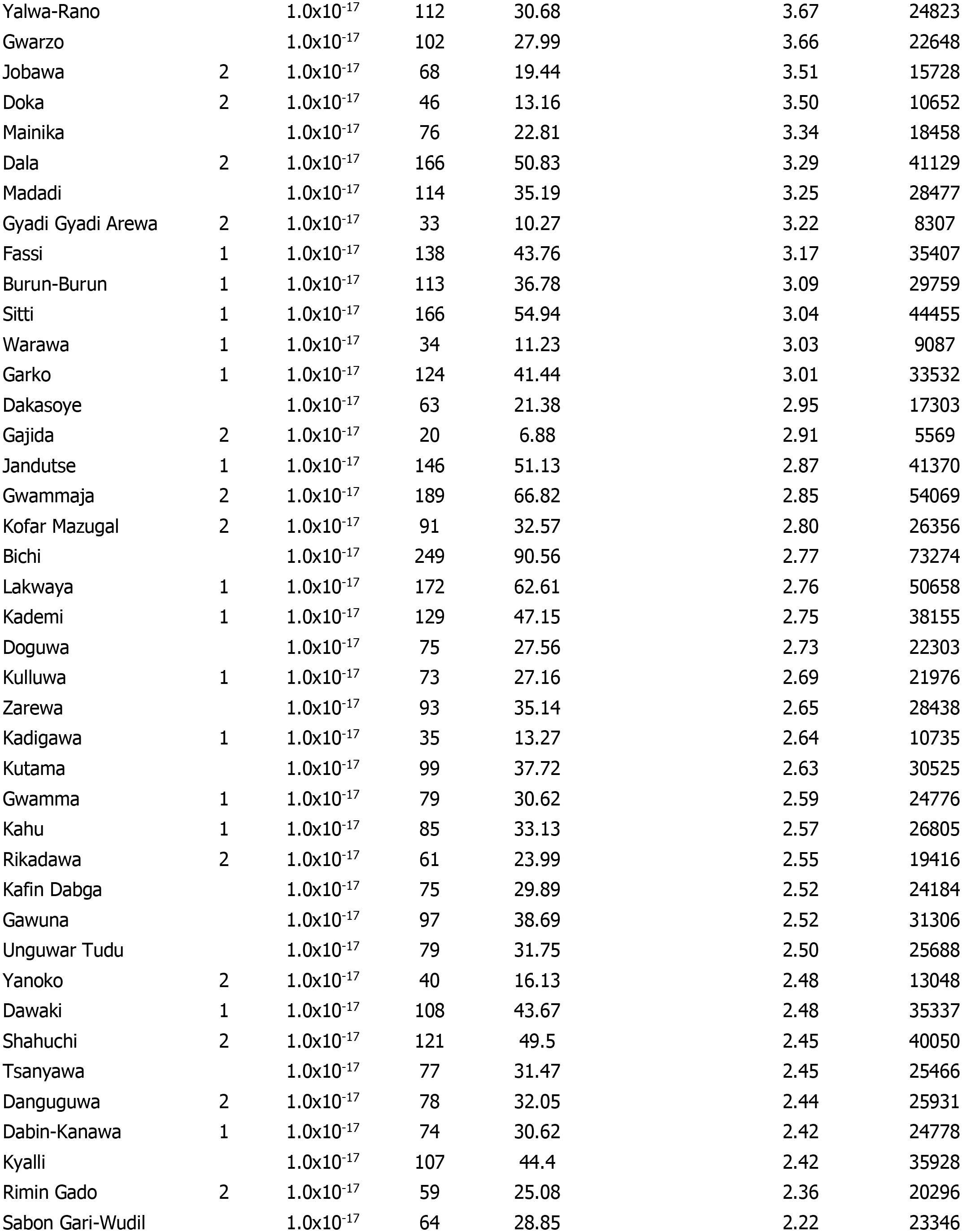
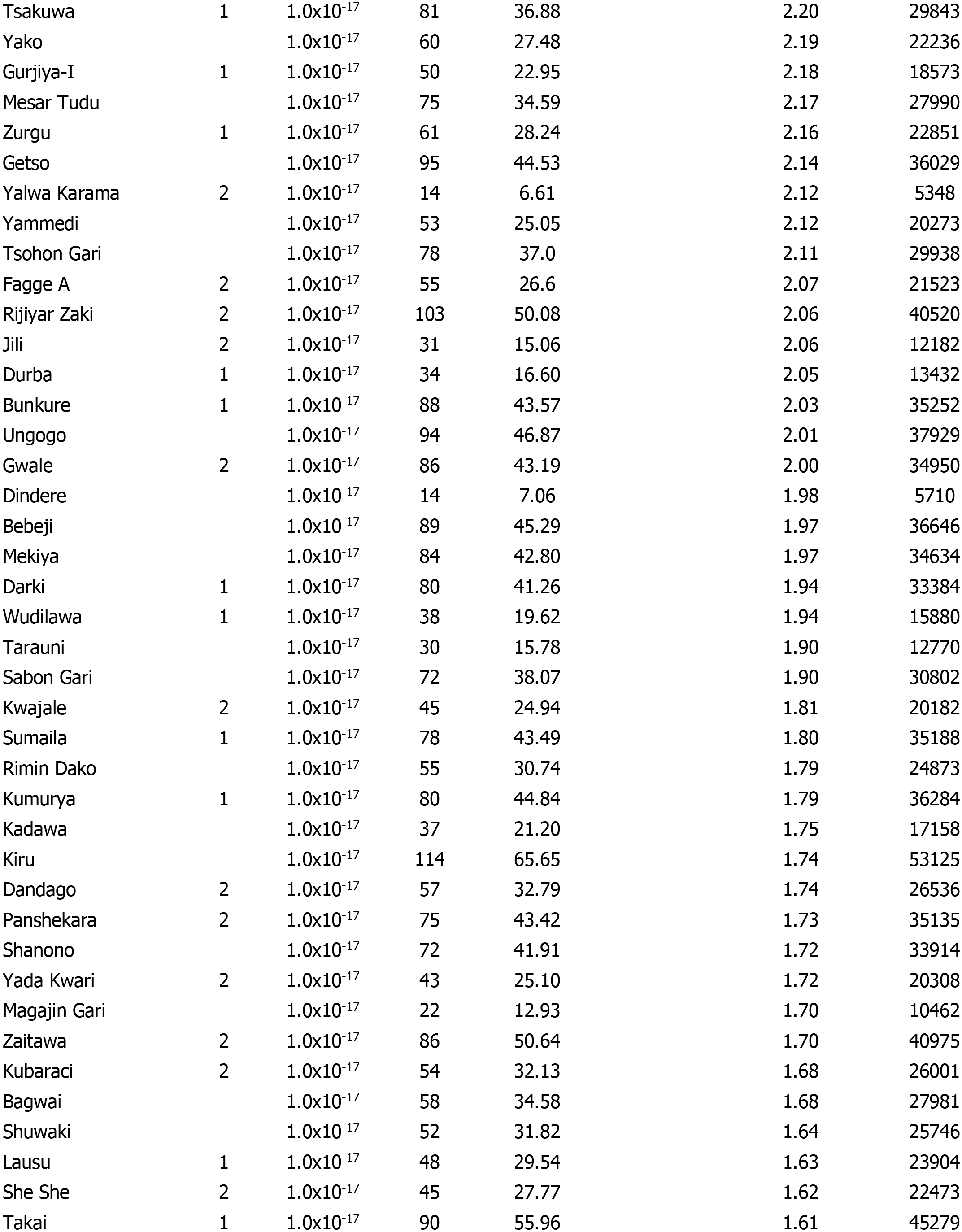
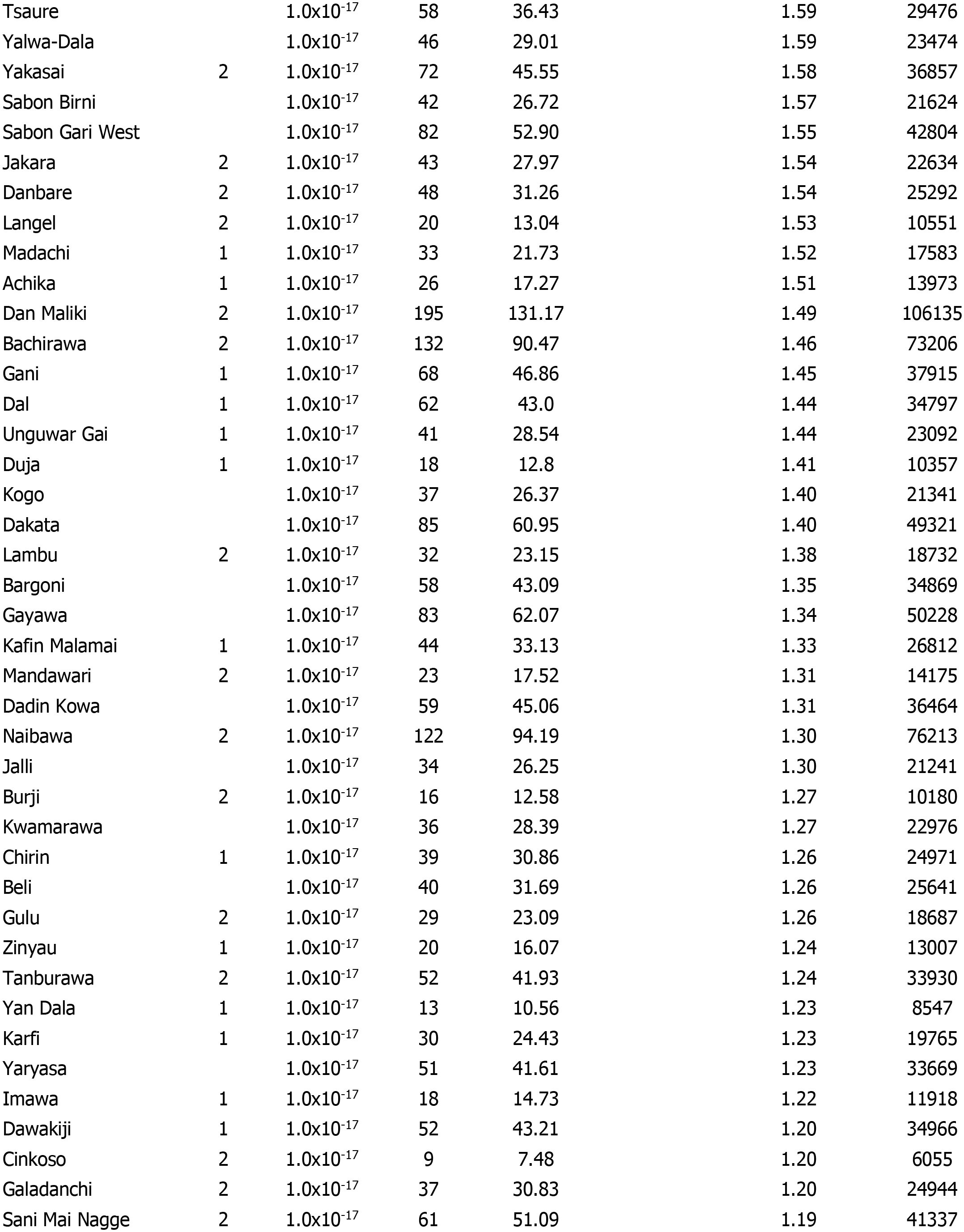
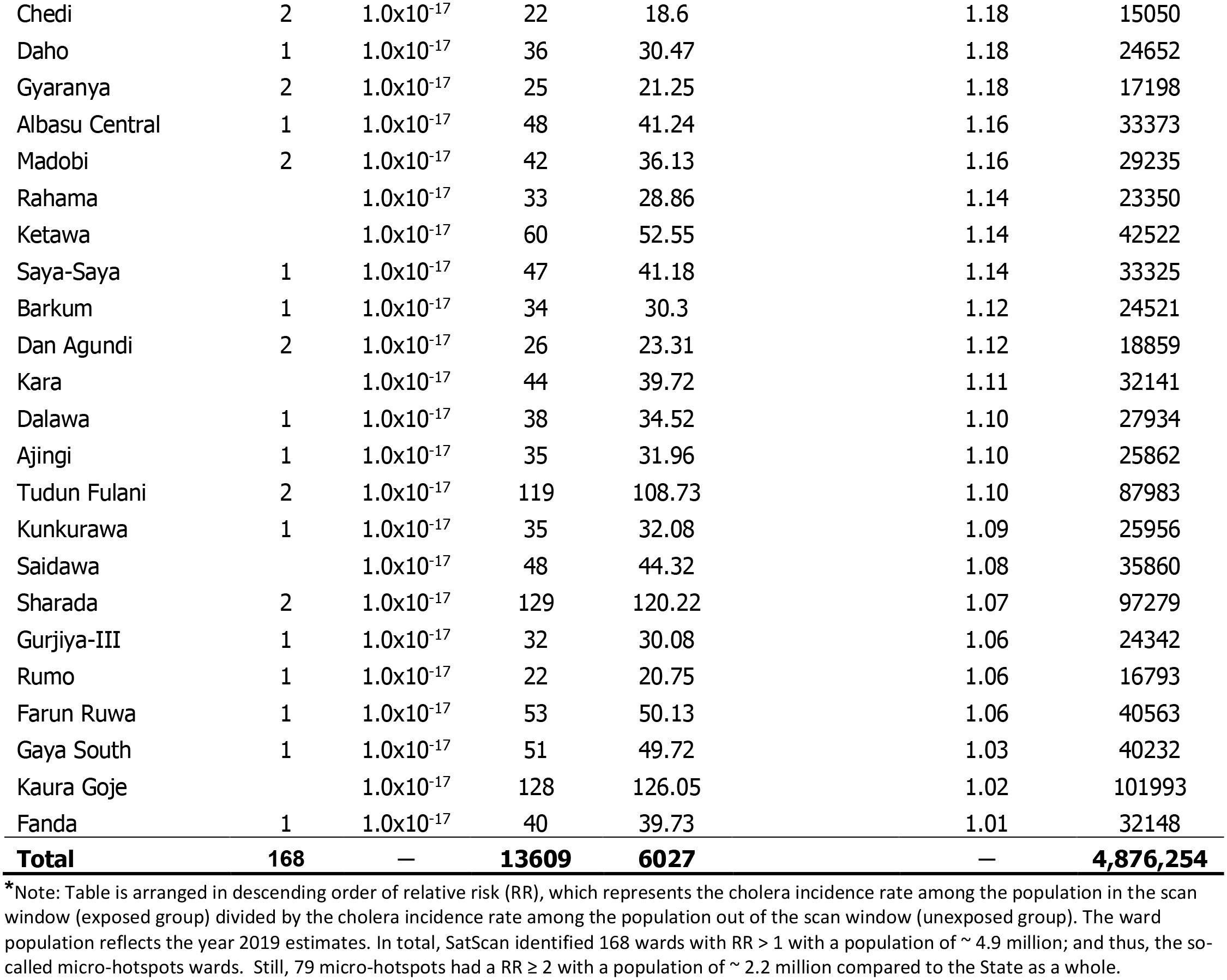
Cholera micro-hotspots classification based on SaTScan (statistical) method.

| Ward | Cluster number | p-value | Observed cases | Expected cases | Relative Risk (RR)* | Population |
| --- | --- | --- | --- | --- | --- | --- |
| Fagge B | 2 | $1.0 \times 10^{-17}$ | 149 | 8.06 | 18.63 | 6525 |
| Kunchi | 3 | $1.0 \times 10^{-17}$ | 310 | 19.96 | 15.79 | 16152 |
| Fagge D2 | 2 | $1.0 \times 10^{-17}$ | 180 | 19.2 | 9.46 | 15539 |
| Gano | 1 | $1.0 \times 10^{-17}$ | 363 | 47.6 | 7.77 | 38513 |
| Tumbau | 1 | $1.0 \times 10^{-17}$ | 246 | 32.11 | 7.75 | 25985 |
| Rijiyar Lemo | 2 | $1.0 \times 10^{-17}$ | 283 | 39.79 | 7.21 | 32200 |
| Gaya North | 1 | $1.0 \times 10^{-17}$ | 315 | 57.49 | 5.56 | 46515 |
| Goron Dutse | 2 | $1.0 \times 10^{-17}$ | 116 | 21.33 | 5.47 | 17259 |
| Kibiya 1 | 1 | $1.0 \times 10^{-17}$ | 98 | 18.68 | 5.27 | 15114 |
| Rano Sabon Gari | 1 | $1.0 \times 10^{-17}$ | 176 | 33.77 | 5.25 | 27324 |
| Sare-Sare | | $1.0 \times 10^{-17}$ | 130 | 26.78 | 4.88 | 21673 |
| Kazurawa | 1 | $1.0 \times 10^{-17}$ | 209 | 44.08 | 4.79 | 35668 |
| Kabo | 2 | $1.0 \times 10^{-17}$ | 159 | 33.85 | 4.73 | 27393 |
| Tofa | 2 | $1.0 \times 10^{-17}$ | 105 | 24.46 | 4.31 | 19795 |
| Kumbotso | 2 | $1.0 \times 10^{-17}$ | 77 | 18.61 | 4.15 | 15060 |
| Utai | 1 | $1.0 \times 10^{-17}$ | 109 | 27.05 | 4.05 | 21892 |
| Wudil | 1 | $1.0 \times 10^{-17}$ | 201 | 53.11 | 3.82 | 42977 |
| Rogo Sabon Gari | | $1.0 \times 10^{-17}$ | 129 | 34.72 | 3.74 | 28096 |
| Magami | 1 | $1.0 \times 10^{-17}$ | 256 | 69.62 | 3.72 | 56336 |
| Yalwan Danziyal | 2 | $1.0 \times 10^{-17}$ | 74 | 20.06 | 3.70 | 16232 |
| Katarkawa | 1 | $1.0 \times 10^{-17}$ | 60 | 16.34 | 3.68 | 13218 |
| Gezawa | | $1.0 \times 10^{-17}$ | 145 | 39.66 | 3.68 | 32091 |
| Yalwa-Rano |  | 1.0x10<sup>-17</sup> | 112 | 30.68 | 3.67 | 24823 |
| Gwarzo |  | 1.0x10<sup>-17</sup> | 102 | 27.99 | 3.66 | 22648 |
| Jobawa | 2 | 1.0x10<sup>-17</sup> | 68 | 19.44 | 3.51 | 15728 |
| Doka | 2 | 1.0x10<sup>-17</sup> | 46 | 13.16 | 3.50 | 10652 |
| Mainika |  | 1.0x10<sup>-17</sup> | 76 | 22.81 | 3.34 | 18458 |
| Dala | 2 | 1.0x10<sup>-17</sup> | 166 | 50.83 | 3.29 | 41129 |
| Madadi |  | 1.0x10<sup>-17</sup> | 114 | 35.19 | 3.25 | 28477 |
| Gyadi Gyadi Arewa | 2 | 1.0x10<sup>-17</sup> | 33 | 10.27 | 3.22 | 8307 |
| Fassi | 1 | 1.0x10<sup>-17</sup> | 138 | 43.76 | 3.17 | 35407 |
| Burun-Burun | 1 | 1.0x10<sup>-17</sup> | 113 | 36.78 | 3.09 | 29759 |
| Sitti | 1 | 1.0x10<sup>-17</sup> | 166 | 54.94 | 3.04 | 44455 |
| Warawa | 1 | 1.0x10<sup>-17</sup> | 34 | 11.23 | 3.03 | 9087 |
| Garko | 1 | 1.0x10<sup>-17</sup> | 124 | 41.44 | 3.01 | 33532 |
| Dakasoye |  | 1.0x10<sup>-17</sup> | 63 | 21.38 | 2.95 | 17303 |
| Gajida | 2 | 1.0x10<sup>-17</sup> | 20 | 6.88 | 2.91 | 5569 |
| Jandutse | 1 | 1.0x10<sup>-17</sup> | 146 | 51.13 | 2.87 | 41370 |
| Gwammaja | 2 | 1.0x10<sup>-17</sup> | 189 | 66.82 | 2.85 | 54069 |
| Kofar Mazugal | 2 | 1.0x10<sup>-17</sup> | 91 | 32.57 | 2.80 | 26356 |
| Bichi |  | 1.0x10<sup>-17</sup> | 249 | 90.56 | 2.77 | 73274 |
| Lakwaya | 1 | 1.0x10<sup>-17</sup> | 172 | 62.61 | 2.76 | 50658 |
| Kademi | 1 | 1.0x10<sup>-17</sup> | 129 | 47.15 | 2.75 | 38155 |
| Doguwa |  | 1.0x10<sup>-17</sup> | 75 | 27.56 | 2.73 | 22303 |
| Kulluwa | 1 | 1.0x10<sup>-17</sup> | 73 | 27.16 | 2.69 | 21976 |
| Zarewa |  | 1.0x10<sup>-17</sup> | 93 | 35.14 | 2.65 | 28438 |
| Kadigawa | 1 | 1.0x10<sup>-17</sup> | 35 | 13.27 | 2.64 | 10735 |
| Kutama |  | 1.0x10<sup>-17</sup> | 99 | 37.72 | 2.63 | 30525 |
| Gwamma | 1 | 1.0x10<sup>-17</sup> | 79 | 30.62 | 2.59 | 24776 |
| Kahu | 1 | 1.0x10<sup>-17</sup> | 85 | 33.13 | 2.57 | 26805 |
| Rikadawa | 2 | 1.0x10<sup>-17</sup> | 61 | 23.99 | 2.55 | 19416 |
| Kafin Dabga |  | 1.0x10<sup>-17</sup> | 75 | 29.89 | 2.52 | 24184 |
| Gawuna |  | 1.0x10<sup>-17</sup> | 97 | 38.69 | 2.52 | 31306 |
| Unguwar Tudu |  | 1.0x10<sup>-17</sup> | 79 | 31.75 | 2.50 | 25688 |
| Yanoko | 2 | 1.0x10<sup>-17</sup> | 40 | 16.13 | 2.48 | 13048 |
| Dawaki | 1 | 1.0x10<sup>-17</sup> | 108 | 43.67 | 2.48 | 35337 |
| Shahuchi | 2 | 1.0x10<sup>-17</sup> | 121 | 49.5 | 2.45 | 40050 |
| Tsanyawa |  | 1.0x10<sup>-17</sup> | 77 | 31.47 | 2.45 | 25466 |
| Danguguwa | 2 | 1.0x10<sup>-17</sup> | 78 | 32.05 | 2.44 | 25931 |
| Dabin-Kanawa | 1 | 1.0x10<sup>-17</sup> | 74 | 30.62 | 2.42 | 24778 |
| Kyalli |  | 1.0x10<sup>-17</sup> | 107 | 44.4 | 2.42 | 35928 |
| Rimin Gado | 2 | 1.0x10<sup>-17</sup> | 59 | 25.08 | 2.36 | 20296 |
| Sabon Gari-Wudil |  | 1.0x10<sup>-17</sup> | 64 | 28.85 | 2.22 | 23346 |
| Tsakuwa | 1 | 1.0x10<sup>-17</sup> | 81 | 36.88 | 2.20 | 29843 |
| Yako |  | 1.0x10<sup>-17</sup> | 60 | 27.48 | 2.19 | 22236 |
| Gurjiya-I | 1 | 1.0x10<sup>-17</sup> | 50 | 22.95 | 2.18 | 18573 |
| Mesar Tudu |  | 1.0x10<sup>-17</sup> | 75 | 34.59 | 2.17 | 27990 |
| Zurgu | 1 | 1.0x10<sup>-17</sup> | 61 | 28.24 | 2.16 | 22851 |
| Getso |  | 1.0x10<sup>-17</sup> | 95 | 44.53 | 2.14 | 36029 |
| Yalwa Karama | 2 | 1.0x10<sup>-17</sup> | 14 | 6.61 | 2.12 | 5348 |
| Yammedi |  | 1.0x10<sup>-17</sup> | 53 | 25.05 | 2.12 | 20273 |
| Tsohon Gari |  | 1.0x10<sup>-17</sup> | 78 | 37.0 | 2.11 | 29938 |
| Fagge A | 2 | 1.0x10<sup>-17</sup> | 55 | 26.6 | 2.07 | 21523 |
| Rijiyar Zaki | 2 | 1.0x10<sup>-17</sup> | 103 | 50.08 | 2.06 | 40520 |
| Jili | 2 | 1.0x10<sup>-17</sup> | 31 | 15.06 | 2.06 | 12182 |
| Durba | 1 | 1.0x10<sup>-17</sup> | 34 | 16.60 | 2.05 | 13432 |
| Bunkure | 1 | 1.0x10<sup>-17</sup> | 88 | 43.57 | 2.03 | 35252 |
| Ungogo |  | 1.0x10<sup>-17</sup> | 94 | 46.87 | 2.01 | 37929 |
| Gwale | 2 | 1.0x10<sup>-17</sup> | 86 | 43.19 | 2.00 | 34950 |
| Dindere |  | 1.0x10<sup>-17</sup> | 14 | 7.06 | 1.98 | 5710 |
| Bebeji |  | 1.0x10<sup>-17</sup> | 89 | 45.29 | 1.97 | 36646 |
| Mekiya |  | 1.0x10<sup>-17</sup> | 84 | 42.80 | 1.97 | 34634 |
| Darki | 1 | 1.0x10<sup>-17</sup> | 80 | 41.26 | 1.94 | 33384 |
| Wudilawa | 1 | 1.0x10<sup>-17</sup> | 38 | 19.62 | 1.94 | 15880 |
| Tarauni |  | 1.0x10<sup>-17</sup> | 30 | 15.78 | 1.90 | 12770 |
| Sabon Gari |  | 1.0x10<sup>-17</sup> | 72 | 38.07 | 1.90 | 30802 |
| Kwajale | 2 | 1.0x10<sup>-17</sup> | 45 | 24.94 | 1.81 | 20182 |
| Sumaila | 1 | 1.0x10<sup>-17</sup> | 78 | 43.49 | 1.80 | 35188 |
| Rimin Dako |  | 1.0x10<sup>-17</sup> | 55 | 30.74 | 1.79 | 24873 |
| Kumurya | 1 | 1.0x10<sup>-17</sup> | 80 | 44.84 | 1.79 | 36284 |
| Kadawa |  | 1.0x10<sup>-17</sup> | 37 | 21.20 | 1.75 | 17158 |
| Kiru |  | 1.0x10<sup>-17</sup> | 114 | 65.65 | 1.74 | 53125 |
| Dandago | 2 | 1.0x10<sup>-17</sup> | 57 | 32.79 | 1.74 | 26536 |
| Panshekara | 2 | 1.0x10<sup>-17</sup> | 75 | 43.42 | 1.73 | 35135 |
| Shanono |  | 1.0x10<sup>-17</sup> | 72 | 41.91 | 1.72 | 33914 |
| Yada Kwari | 2 | 1.0x10<sup>-17</sup> | 43 | 25.10 | 1.72 | 20308 |
| Magajin Gari |  | 1.0x10<sup>-17</sup> | 22 | 12.93 | 1.70 | 10462 |
| Zaitawa | 2 | 1.0x10<sup>-17</sup> | 86 | 50.64 | 1.70 | 40975 |
| Kubaraci | 2 | 1.0x10<sup>-17</sup> | 54 | 32.13 | 1.68 | 26001 |
| Bagwai |  | 1.0x10<sup>-17</sup> | 58 | 34.58 | 1.68 | 27981 |
| Shuwaki |  | 1.0x10<sup>-17</sup> | 52 | 31.82 | 1.64 | 25746 |
| Lausu | 1 | 1.0x10<sup>-17</sup> | 48 | 29.54 | 1.63 | 23904 |
| She She | 2 | 1.0x10<sup>-17</sup> | 45 | 27.77 | 1.62 | 22473 |
| Takai | 1 | 1.0x10<sup>-17</sup> | 90 | 55.96 | 1.61 | 45279 |
| Tsaure |  | 1.0x10<sup>-17</sup> | 58 | 36.43 | 1.59 | 29476 |
| Yalwa-Dala |  | 1.0x10<sup>-17</sup> | 46 | 29.01 | 1.59 | 23474 |
| Yakasai | 2 | 1.0x10<sup>-17</sup> | 72 | 45.55 | 1.58 | 36857 |
| Sabon Birni |  | 1.0x10<sup>-17</sup> | 42 | 26.72 | 1.57 | 21624 |
| Sabon Gari West |  | 1.0x10<sup>-17</sup> | 82 | 52.90 | 1.55 | 42804 |
| Jakara | 2 | 1.0x10<sup>-17</sup> | 43 | 27.97 | 1.54 | 22634 |
| Danbare | 2 | 1.0x10<sup>-17</sup> | 48 | 31.26 | 1.54 | 25292 |
| Langel | 2 | 1.0x10<sup>-17</sup> | 20 | 13.04 | 1.53 | 10551 |
| Madachi | 1 | 1.0x10<sup>-17</sup> | 33 | 21.73 | 1.52 | 17583 |
| Achika | 1 | 1.0x10<sup>-17</sup> | 26 | 17.27 | 1.51 | 13973 |
| Dan Maliki | 2 | 1.0x10<sup>-17</sup> | 195 | 131.17 | 1.49 | 106135 |
| Bachirawa | 2 | 1.0x10<sup>-17</sup> | 132 | 90.47 | 1.46 | 73206 |
| Gani | 1 | 1.0x10<sup>-17</sup> | 68 | 46.86 | 1.45 | 37915 |
| Dal | 1 | 1.0x10<sup>-17</sup> | 62 | 43.0 | 1.44 | 34797 |
| Unguwar Gai | 1 | 1.0x10<sup>-17</sup> | 41 | 28.54 | 1.44 | 23092 |
| Duja | 1 | 1.0x10<sup>-17</sup> | 18 | 12.8 | 1.41 | 10357 |
| Kogo |  | 1.0x10<sup>-17</sup> | 37 | 26.37 | 1.40 | 21341 |
| Dakata |  | 1.0x10<sup>-17</sup> | 85 | 60.95 | 1.40 | 49321 |
| Lambu | 2 | 1.0x10<sup>-17</sup> | 32 | 23.15 | 1.38 | 18732 |
| Bargoni |  | 1.0x10<sup>-17</sup> | 58 | 43.09 | 1.35 | 34869 |
| Gayawa |  | 1.0x10<sup>-17</sup> | 83 | 62.07 | 1.34 | 50228 |
| Kafin Malamai | 1 | 1.0x10<sup>-17</sup> | 44 | 33.13 | 1.33 | 26812 |
| Mandawari | 2 | 1.0x10<sup>-17</sup> | 23 | 17.52 | 1.31 | 14175 |
| Dadin Kowa |  | 1.0x10<sup>-17</sup> | 59 | 45.06 | 1.31 | 36464 |
| Naibawa | 2 | 1.0x10<sup>-17</sup> | 122 | 94.19 | 1.30 | 76213 |
| Jalli |  | 1.0x10<sup>-17</sup> | 34 | 26.25 | 1.30 | 21241 |
| Burji | 2 | 1.0x10<sup>-17</sup> | 16 | 12.58 | 1.27 | 10180 |
| Kwamarawa |  | 1.0x10<sup>-17</sup> | 36 | 28.39 | 1.27 | 22976 |
| Chirin | 1 | 1.0x10<sup>-17</sup> | 39 | 30.86 | 1.26 | 24971 |
| Beli |  | 1.0x10<sup>-17</sup> | 40 | 31.69 | 1.26 | 25641 |
| Gulu | 2 | 1.0x10<sup>-17</sup> | 29 | 23.09 | 1.26 | 18687 |
| Zinyau | 1 | 1.0x10<sup>-17</sup> | 20 | 16.07 | 1.24 | 13007 |
| Tanburawa | 2 | 1.0x10<sup>-17</sup> | 52 | 41.93 | 1.24 | 33930 |
| Yan Dala | 1 | 1.0x10<sup>-17</sup> | 13 | 10.56 | 1.23 | 8547 |
| Karfi | 1 | 1.0x10<sup>-17</sup> | 30 | 24.43 | 1.23 | 19765 |
| Yaryasa |  | 1.0x10<sup>-17</sup> | 51 | 41.61 | 1.23 | 33669 |
| Imawa | 1 | 1.0x10<sup>-17</sup> | 18 | 14.73 | 1.22 | 11918 |
| Dawakiji | 1 | 1.0x10<sup>-17</sup> | 52 | 43.21 | 1.20 | 34966 |
| Cinkoso | 2 | 1.0x10<sup>-17</sup> | 9 | 7.48 | 1.20 | 6055 |
| Galadanchi | 2 | 1.0x10<sup>-17</sup> | 37 | 30.83 | 1.20 | 24944 |
| Sani Mai Nagge | 2 | 1.0x10<sup>-17</sup> | 61 | 51.09 | 1.19 | 41337 |
| Chedi | 2 | 1.0x10<sup>-17</sup> | 22 | 18.6 | 1.18 | 15050 |
| Daho | 1 | 1.0x10<sup>-17</sup> | 36 | 30.47 | 1.18 | 24652 |
| Gyaranya | 2 | 1.0x10<sup>-17</sup> | 25 | 21.25 | 1.18 | 17198 |
| Albasu Central | 1 | 1.0x10<sup>-17</sup> | 48 | 41.24 | 1.16 | 33373 |
| Madobi | 2 | 1.0x10<sup>-17</sup> | 42 | 36.13 | 1.16 | 29235 |
| Rahama |  | 1.0x10<sup>-17</sup> | 33 | 28.86 | 1.14 | 23350 |
| Ketawa |  | 1.0x10<sup>-17</sup> | 60 | 52.55 | 1.14 | 42522 |
| Saya-Saya | 1 | 1.0x10<sup>-17</sup> | 47 | 41.18 | 1.14 | 33325 |
| Barkum | 1 | 1.0x10<sup>-17</sup> | 34 | 30.3 | 1.12 | 24521 |
| Dan Agundi | 2 | 1.0x10<sup>-17</sup> | 26 | 23.31 | 1.12 | 18859 |
| Kara |  | 1.0x10<sup>-17</sup> | 44 | 39.72 | 1.11 | 32141 |
| Dalawa | 1 | 1.0x10<sup>-17</sup> | 38 | 34.52 | 1.10 | 27934 |
| Ajingi | 1 | 1.0x10<sup>-17</sup> | 35 | 31.96 | 1.10 | 25862 |
| Tudun Fulani | 2 | 1.0x10<sup>-17</sup> | 119 | 108.73 | 1.10 | 87983 |
| Kunkurawa | 1 | 1.0x10<sup>-17</sup> | 35 | 32.08 | 1.09 | 25956 |
| Saidawa |  | 1.0x10<sup>-17</sup> | 48 | 44.32 | 1.08 | 35860 |
| Sharada | 2 | 1.0x10<sup>-17</sup> | 129 | 120.22 | 1.07 | 97279 |
| Gurjiya-III | 1 | 1.0x10<sup>-17</sup> | 32 | 30.08 | 1.06 | 24342 |
| Rumo | 1 | 1.0x10<sup>-17</sup> | 22 | 20.75 | 1.06 | 16793 |
| Farun Ruwa | 1 | 1.0x10<sup>-17</sup> | 53 | 50.13 | 1.06 | 40563 |
| Gaya South | 1 | 1.0x10<sup>-17</sup> | 51 | 49.72 | 1.03 | 40232 |
| Kaura Goje |  | 1.0x10<sup>-17</sup> | 128 | 126.05 | 1.02 | 101993 |
| Fanda | 1 | 1.0x10<sup>-17</sup> | 40 | 39.73 | 1.01 | 32148 |
| **Total** | **168** | **—** | **13609** | **6027** | **—** | **4,876,254** |

\*Note: Table is arranged in descending order of relative risk (RR), which represents the cholera incidence rate among the population in the scan window (exposed group) divided by the cholera incidence rate among the population out of the scan window (unexposed group). The ward population reflects the year 2019 estimates. In total, SatScan identified 168 wards with RR > 1 with a population of ~ 4.9 million; and thus, the so-called micro-hotspots wards. Still, 79 micro-hotspots had a RR ≥ 2 with a population of ~ 2.2 million compared to the State as a whole.

#### Micro-hotspots based on the GTFCC method

Between 2010 and 2019, the top 10 wards with the highest mean annual incidence rates of cholera were evenly split between urban and rural locations (five in each location (S3 Fig)). In the ten years of the study period, cholera occurred in 89.5% (433/484) of all the wards, and of these, 82.2% (356/433) were rural wards. The mean annual incidence rate for each ward that reported cholera ranged from 0.27 in Kachako to 310.68 per 100,000 persons in Fagge B (S3 Fig). Based on the 55th and 71.6th percentiles, the cut-off for the mean annual incidence rate and persistence were 10/100,000 persons and 2.50%, respectively. Application of these criteria yielded 115 wards as T1 micro-hotspot, 105 as T2 micro-hotspot, 23 as T3 micro-hotspot, and 241 as T4 micro-hotspot (Table 4, S1 Table). In Fig 7, we present the cholera micro-hotspot classification chart (Fig 7A) and map (Fig 7B) while Table 4 and S1 Table provide the list of the wards under the different micro-hotspots categories. The populations with these micro-hotspot prioritizations include 4,505,370 for T1, 2,413,291 for T2, 1,570,479 for T3, and 5,811,246 for T4 (Table 4, S1 Table).

**Fig 7.**
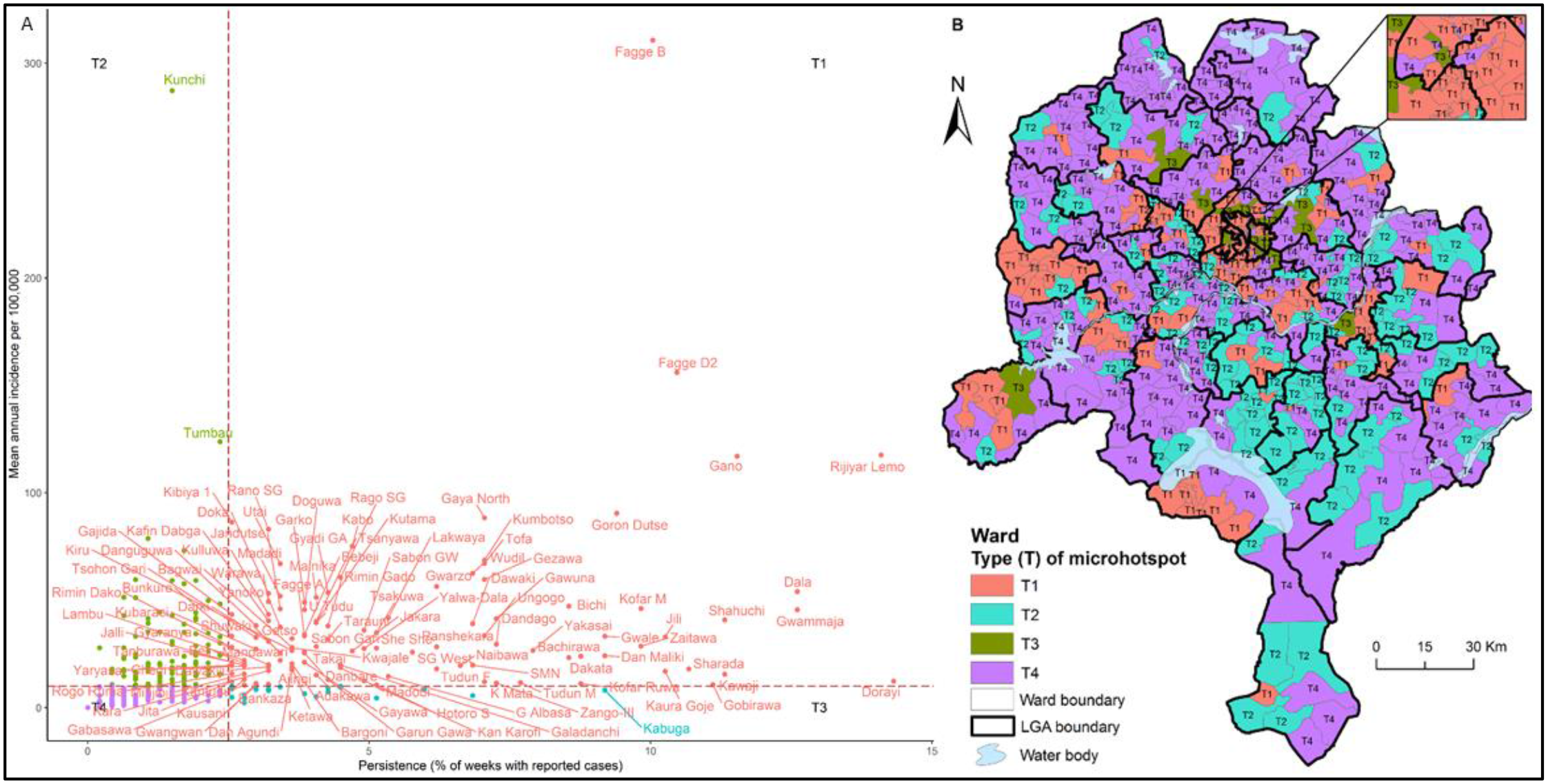
Cholera micro-hotspot classification chart and map for Kano State following GTFCC method, 2010-2019. (A) The persistence cut-off point is 2.5% and the mean annual incidence rate cut-off point is 10/100,000 population. T represents types of micro-hotspots. T1 (high priority) reflects high persistence and mean annual incidence; T2 (medium priority) shows high persistence and low mean annual incidence; T3 (medium priority) indicates low persistence and high mean annual incidence; and T4 (low priority) depicts low persistence and mean annual incidence, respectively. The dots denote wards in the four types of micro-hotspot quadrants. The figure shows 115 (T1), 105 (T2), 23 (T3), and 241 (T4) risk types. Compared with the statistical map in Fig 4, this GTFCC method misses 50 micro-hotspot wards in the T1 quadrant. However, this could be the result of the cut-off point. Note that Fig 7 show labels for all T1 micro-hotdpots, but only two for T2 (Kunchi and Tumbau), one for T3 (Kabuga), and none for T4, as space limitattion did not permit showing all labels. (B) Spatial map of the T (types of micro-hotspots) in relation to inland water bodies.

**Table 4.**
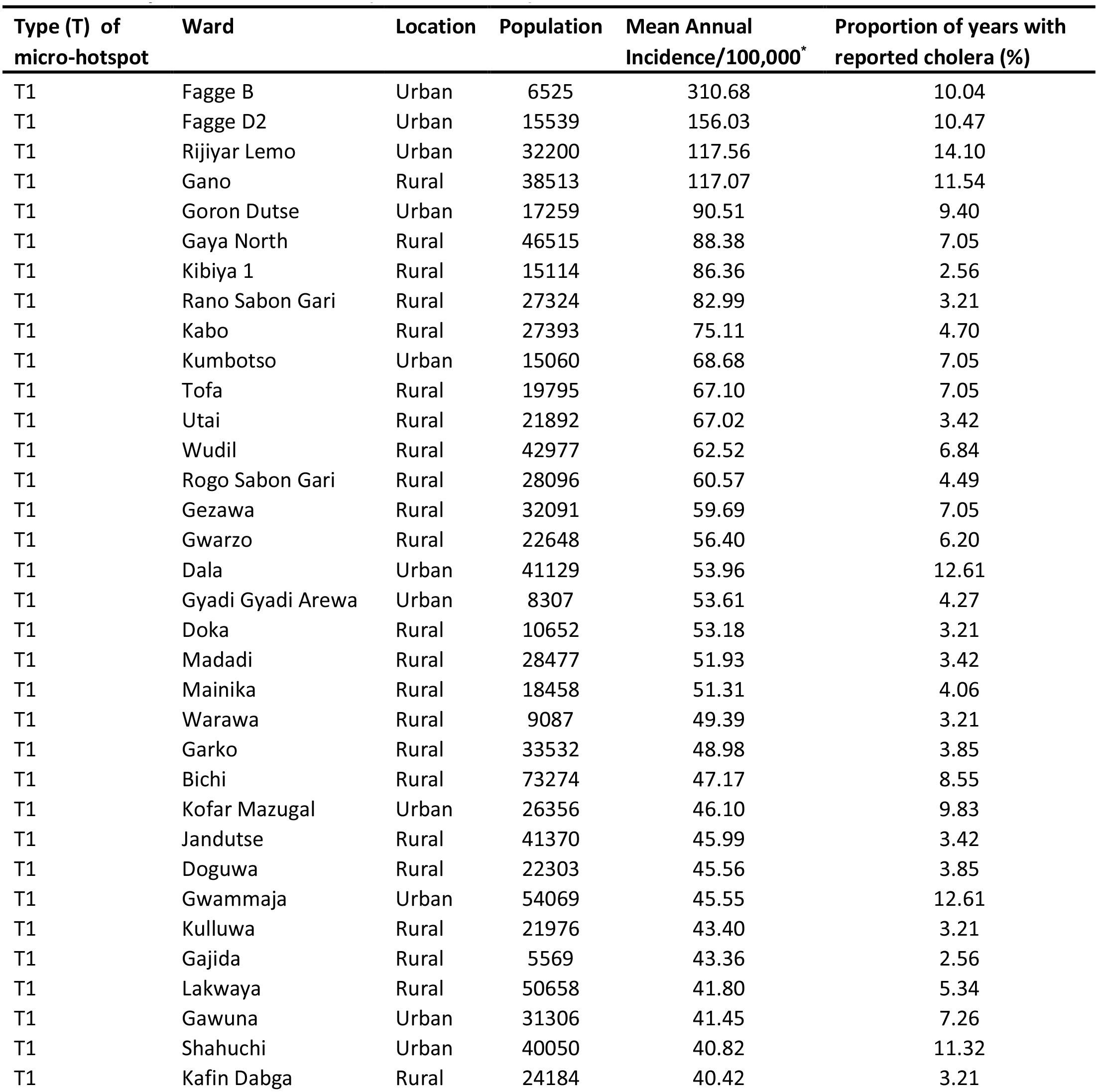
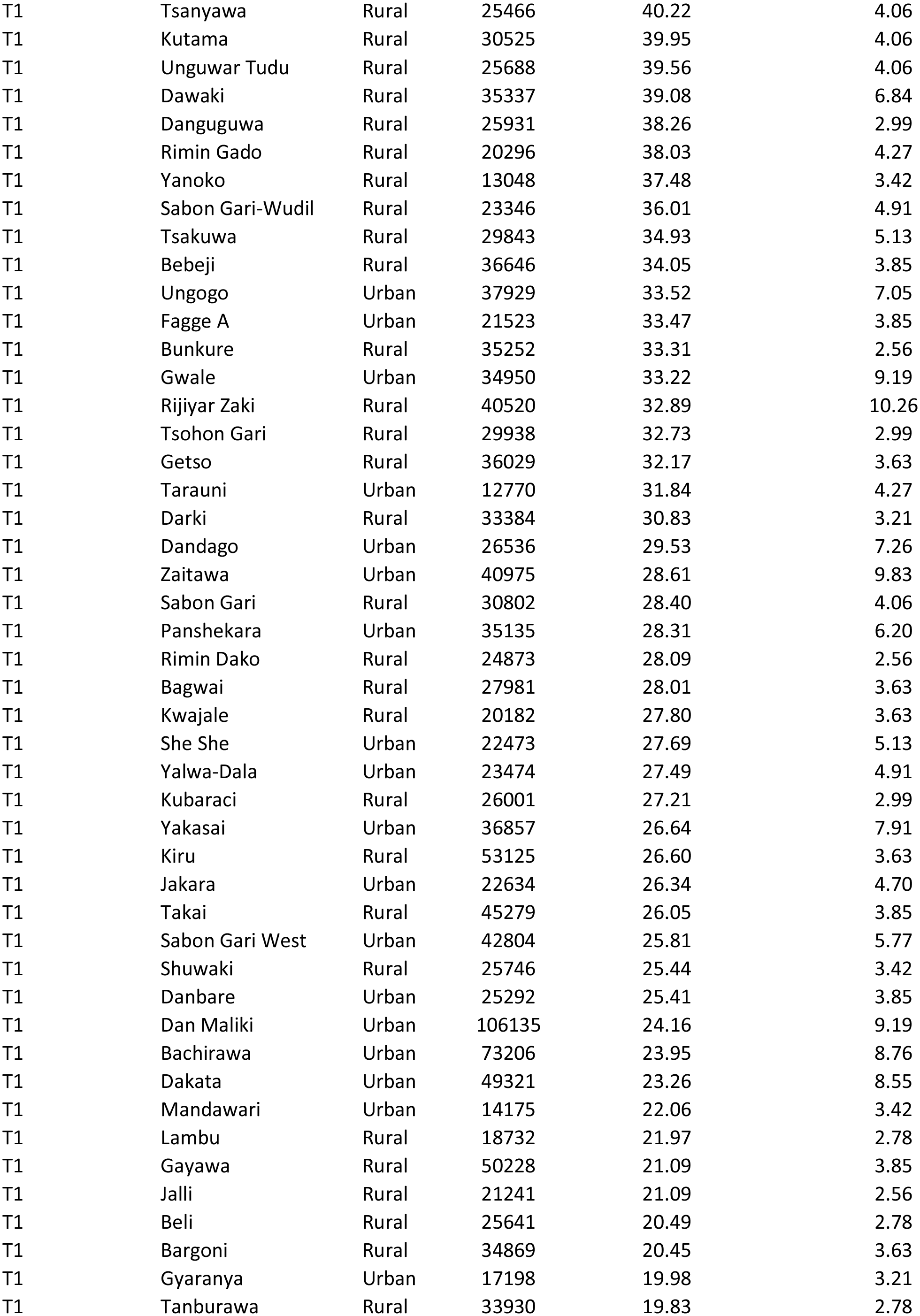
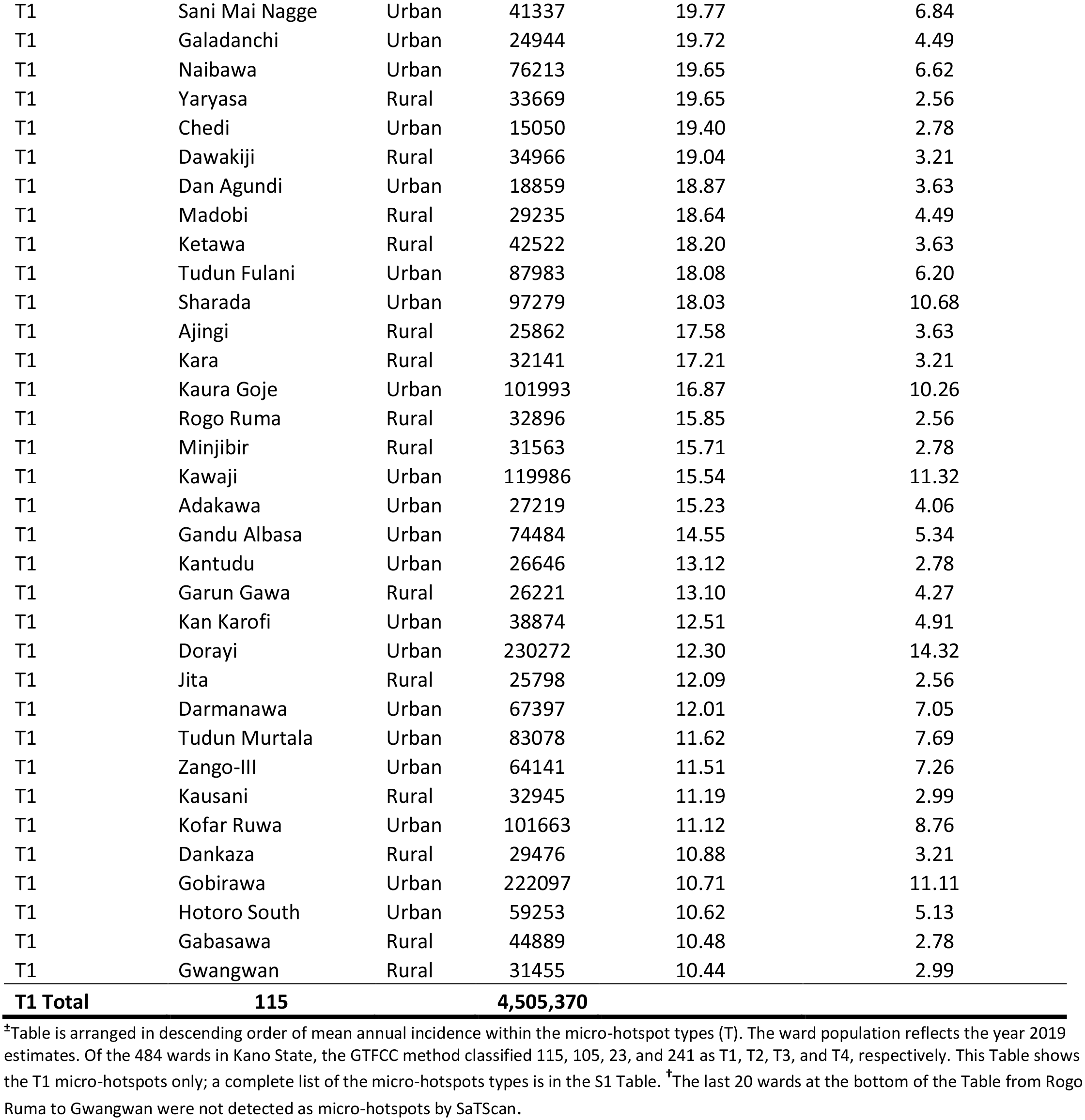
Wards with the type of micro-hotspots based on mean annual incidence rate and proportion of weeks with reported cholera cases (GTFCC method)

In keeping with our objective of comparing SaTScan and GTFCC methods in analyzing micro-hotspot patterns, both showed points of similarities and dissimilarities. Remarkably, for 2010-2019, both methods agreed on eight out of the top ten ranked micro-hotspots. Yet, SaTScan identified Kunchi and Tumbau within its top ten ranked micro-hotspots while GTFCC classified Kabo and Kumbotso within its top ten ranked micro-hotspots (Tables 3 and 4). Similarly, considering the T1 and some T2 wards, of the 168 micro-hotspots detected using SaTScan, 57.74% (97/168) and 42.26% (71/168) were classified as T1 and T2 by GTFCC method, respectively. Conversely, the GTFCC tool classified 20 wards from Rogo Ruma to Gwangwan as T1 (Table 4) when SaTScan did not detect these wards as micro-hotspots; i.e., the 20 wards had a RR<1. Finally, 52.98% (89/168) of micro-hotspots detected by SaTScan had a RR < 2 (Table T3); nonetheless, GTFCC classified most of these wards as T1 (Table 4). For instance, according to SaTScan, the Kaura Goje micro-hotspot has a RR = 1.02 (Table 3), but was a T1 micro-hotspot according to GTFCC with a very high cholera persistence of 10.26% (Table 4).

## Discussion

The goals of this study were to provide an analysis of the Spatio-temporal epidemiology of cholera in Kano State, Nigeria, and to describe alternative methods for identifying micro-hotspots for cholera. Identifying these hotspots is essential for controlling cholera to develop interventions at a local community (ward level) and is needed for the implementation of the Nigeria NSPACC. Thus, our findings on the clinical, demographic, and behavioral characteristics, as well as geographic distributions, space-time patterns of disease clusters, and micro-hotspots using data from 2010 to 2019, have public health implications.

Between 2010 and 2019, we found that location, season, gender, and age significantly affected the risk of cholera in Kano State. Cholera in Kano State occurs in both urban and rural locations/settings with only minor differences in rates of infection in the two settings. The disease occurs in the dry and rainy seasons alike, but intense weekly transmission occurs during the rainy season with peaks in August. The spatial distribution of cholera showed a considerable heterogeneous pattern across the state, but with large space-time clusters at the Center and eastern portions between 2013 and 2014. We detected 168 micro-hotspots, and the two methods we used generally agreed. Amongst women, cholera survival significantly depended on the setting (*p-value* = 0.003). The age distribution of cases and deaths found higher rates among those 15 years and younger (Fig 2). The CFR was higher in rural settings (3.4%) but decreased with increasing age in both settings. We observed a 77.4% case severity in the urban compared to 53.4% in the rural setting. For care-seeking behaviour, 52.1% of cases in the urban setting sought clinical care on the same day of symptom onset compared with 82.7% of their peers in the rural location. Overall, confirmation of the diagnosis using RDTs was very low (1.3%), especially in a rural location (1.0%).

Within the urban-rural divide of cholera in Kano State, rural populations had more cases and deaths than urban dwellers, especially rural women. Though the rural population is more than two and a half times (2.6) that of the urban, the AR between the two differs by only 10.8 per 100000 inhabitants (Table 1); the cholera mortality rate is slightly higher in the rural than urban areas. While cholera has mostly been thought to be higher in urban areas [46–51] and lower in the rural than crowded urban slums [50, 52], our finding that cholera risk is not significantly higher in the urban setting of Kano State is consistent with findings of high cholera risk in rural settings in Guinea-Bissau [53], Zimbabwe [54], and Haiti [54]. The finding that women and children in the rural setting had more cases of cholera than women and children in the urban setting (Fig. 2) could be attributed to rural practices regarding food (eating habits), hygiene, and gatherings such as funerals, weddings, markets, and the use of surface water. Further, children might be finishing adults’ plates, women preparing food using surface water, and attending and eating at funerals/weddings without proper hand hygiene, all factors that have been associated with cholera outbreaks in Cameroon [55] and Bangladesh [56].

For 2010 to 2019, we confirmed that in Kano State, though the disease occurs in the dry and rainy seasons alike, higher rates of cholera occurred during the rainy season (June to September) with a peak in August (Fig. 2) [9–12]. Most importantly, cases in the rural and urban settings peak asynchronously, i.e., when cases in the rural setting are at a peak, those in the urban location are at a trough (Fig 3). Control efforts would benefit from studies of the seasonality of cholera in Kano State that correlate rainfall data with cases of cholera.

We observed that CFRs are high (3.5% rural vs 3.0% urban), far exceeds the <1% recommended by the WHO and decreasing with increasing age (Table 1), as has been documented elsewhere [57]. High CFRs call for better hospital management of cases in general, but with particular attention on children <5 years old, and improvements in surveillance to trigger a timely response. The decreasing rates of disease with age are thought to be related to acquired immunity, and this is supported by the increasingly elevated vibriocidal antibody titers by age in the population [58, 59]. For 2010 and 2019, more than three-quarters of cases in the urban setting were severe, however just over half of them sought care on the same day of cholera onset. The reverse is true in the rural setting; slightly more than half of the cases in the rural location were judged to be severe, and nearly three-quarters of them sought care on the same day of disease onset. The reasons for higher cholera severity and poorer care-seeking behavior in the urban setting are not well understood. Likewise, reasons for lower severity, though still high, and better care-seeking behavior in a rural setting are not well understood. This could be related to educational status, income, occupation, and body mass index [60]. Perhaps high mobility and lack of time to seek timely care in an urban setting could be contributing factors. Conceivably, the rural population may be less mobile and have more discretionary time to seek immediate care at health facilities.

The present study also shows that between 2010 and 2019, sample collection and testing to ascertain *V. cholerae* in stool was only 1.3% overall. Likely, the 2.2% to 1.0% (reflected in the RDT positivity rates 61.7% to 63.8%) disparity in urban and rural stool sample testing indicates the difficulty in reaching rural locations with RDTs. Inadequate testing occurred among <5 years and 5-14 years olds in the rural setting, age groups in which suspected cases have a higher chance of not being cholera. Unfortunately, data on culture confirmation was not available for analysis. This insufficient testing calls for urgent research to understand the limiting factors to laboratory surveillance including, but not limited to, personnel and availability of RDT kits in the urban and rural settings alike.

There was a clear tendency for wards reporting cholera to be clustered around inland water bodies of the northern, central, and southeastern portions of the state (Fig 5). We detected 168 wards that were at high risk of cholera and were considered to be micro-hotspots, compared to other wards, and both methods used were generally in agreement.

However, SaTScan did not detect the bottom 20 T1 micro-hotspots of Table 4 (from Rogo Ruma to Gwangwan) as micro-hotspots. SaTScan is dependent primarily on incidence whereas the GTFCC method includes both incidence and persistence. Thus, areas with higher incidence may be better identified when using the GTFCC method. Yet, as both methods generally agreed (Figs 6 and 7), our finding corroborates our hypothesis that although an LGA may be detected as a hotspot LGA not all wards within it are micro-hotspots [8]. For example, in our earlier hotspot detection at the LGA level, Rogo LGA was not classified as hotspot LGA [8], but at the fine-scale of ward, both methods detected 3-4 micro-hotspots in Rogo (Figs 6 and 7). Also, Kunchi LGA was classified as hotspot LGA, but only one ward in Kunchi LGA is a micro-hotspot, and so forth. Therefore, we conclude that hotspot maps at the ward level, not hotspot maps at the LGA level, are best suited to target cholera interventions including vaccines.

As our main objective was to provide NSPACC with data to guide interventions, the micro-hotpots in Table 3 would benefit most from priority vaccination by the National Primary Health Care Development Agency, Kano State Ministry of Health, and NCDC while longer-term WASH infrastructure is being put in place in line with the global roadmap of ending cholera by 2030. As NSPACC plans to provide vaccines in Kano State between 2020 and 2023 this analysis can support their preparation and specific plans for vaccination. The population of wards with a significantly higher risk was 4,876,254 while the population of the wards with a relative risk ≥2 was 2,119,974. SaTScan has been widely used and found very helpful in detecting cholera clusters (hotspots) to inform interventions in Cameroon [17], Uganda [18], Zambia [46], and Tanzania [47]. By contrast, the GTFCC tool is new, and excepting Ngwa *et al*. [8], we did not find it being used in peer review published reports. Therefore, more scholarly work is needed to illuminate the viability of the GTFCC tool for use by local policymakers who will need to include local factors with hotspot analysis when making priorities for interventions.

This study has important limitations as well as strengths. Testing of suspected cases was very low. As such, we cannot roll out misclassification of cases based on clinical criteria leading to underestimation or overestimation of the true rates of cholera. Although unlikely, retrospective determination of ward of cases could have led to assigning cases to the wrong ward. Nonetheless, the use of the health facility-based line-list data spanning ten years afforded the prospect to describe demographic, testing, and behavioral characteristics of cases as well as patterns of spatial distribution, Spatio-temporal clusters, and micro-hotspots needed to inform intervention including vaccine; and thus, constitute major strengths of the study. Other strengths include illuminating urban/rural disparity in cholera cases and deaths besides comparing two methods to micro-hotspots detection, which generally agreed. Although SaTScan is widely used, its use demands a strong statistical knowledge compared to the GTFCC method, which requires only entering data into Microsoft Excel. As such, the later method can readily be applied in resource-poor settings without statisticians for hotspots identification. Finally, policymakers have been served with analysis stratified by gender, age, and location leading to uncovering the rural-urban disparity in cholera transmission.

In sum, in 2017, the road map to ending cholera by 2030 called for focusing interventions in cholera hotspots [15]. In 2018, Nigeria was determined to pre-emptively vaccinate high-risk populations in identified hotspots nationally between 2018 and 2023 [16]. We conclude that hotspot maps at the ward level, not hotspot maps at the LGA level, are best suited to target cholera interventions including vaccines. This study has served the NSPACC with the high-priority micro-hotspots of cholera in Kano State to target interventions including vaccines towards cholera elimination from the state by 2030.

## Supporting information

Supplement_files_v3_medrxiv

## Data Availability

Data cannot be shared publicly because of concerns that some wards have very small populations and individuals might be identified. Data are available from the Nigeria Centre for Disease Control or Kano Ministry of Health via Drs. Chikwe Ihekweazu
and/or 'Imam Bello'
for researchers who meet the criteria for access to confidential data.

