## Supplement_files_v3_medrxiv for "The micro-hotspots of cholera in Kano State, Nigeria, 2010-2019—analysis of patient characteristics, Spatio-temporal patterns and contextual determinants at the ward level"

Supplements.

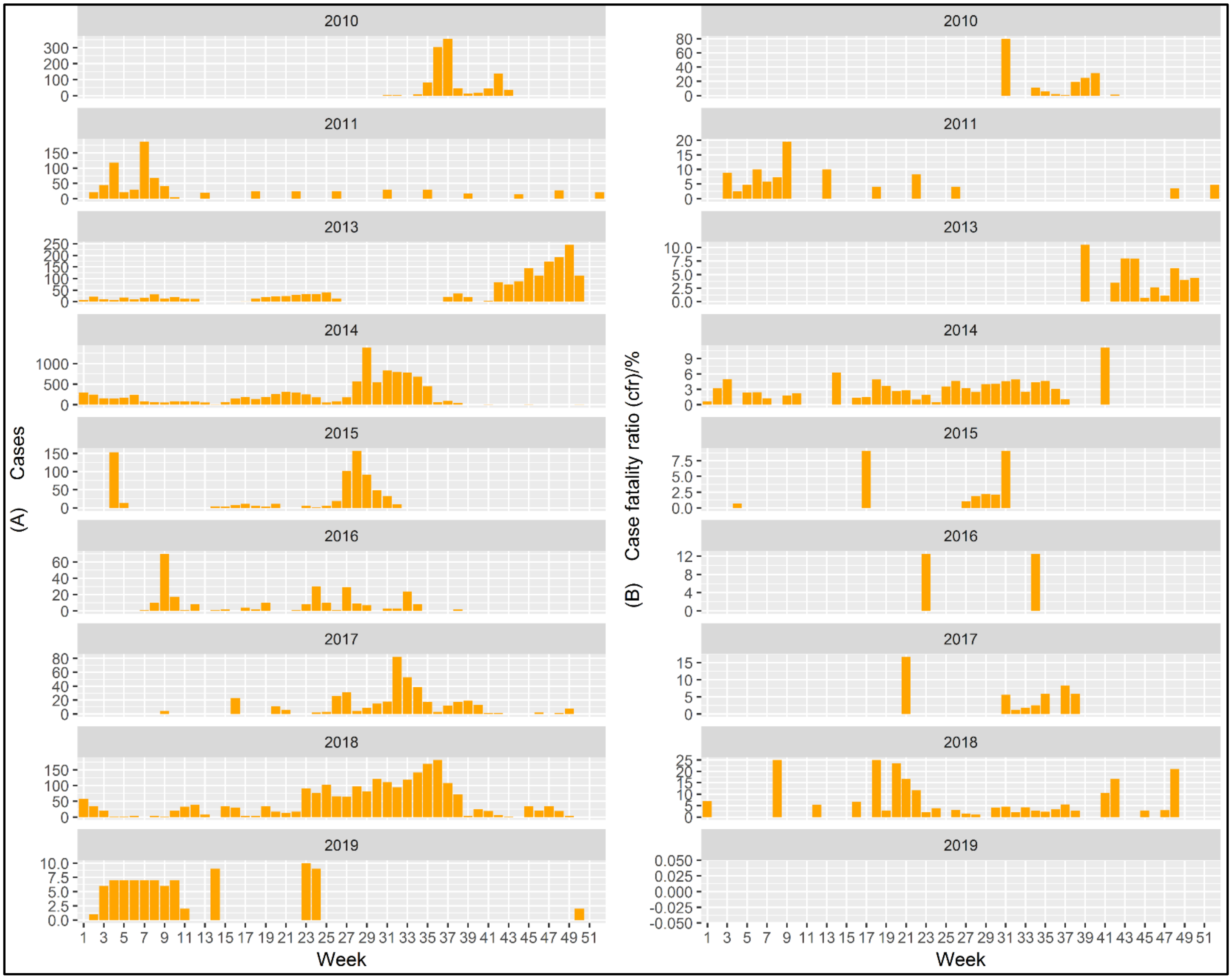


**S1 Fig. Weekly occurrence of cholera epidemics in Kano State, Nigeria, 2010-2019.** (A) Reported cases and (B) case fatality ratio (CFR) are shown on the Y-axis while the X-axis represent the epidemiological week of disease onset. Reported outbreaks in this time period started in week 31 of 2010 (A) with 80% case fatality ratio (CFR) (B) representing the worst CFR in the entire study period. (A) In subsequent years, the disease occurred with multiple waves and peaks through the year with 2014 reporting over a 1300 cases in week 27 alone. Weekly CFRs far exceeded below 1% as recommended by the WHO, although there were no fatalities in 2019 (B). No cholera epidemic was reported in 2012.

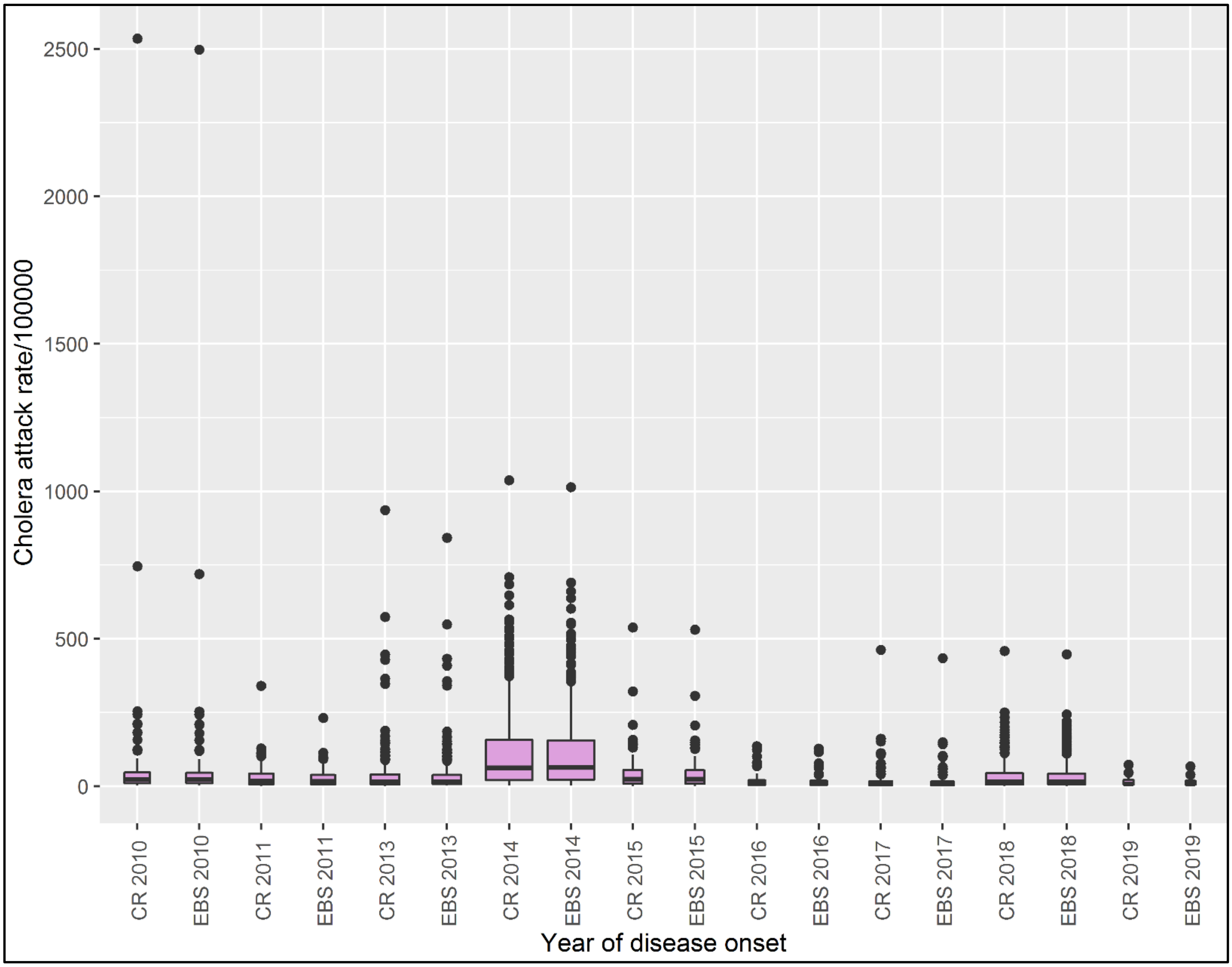


S2 Fig. Comparative boxplots for crude and Empirical Bayes Smoothed attack rates per 100,000 inhabitants. Plot shows minor differences between unsmoothed crude rates (CR) and Smoothed (EBS) rates; and thus, CR rates were used in analysis.

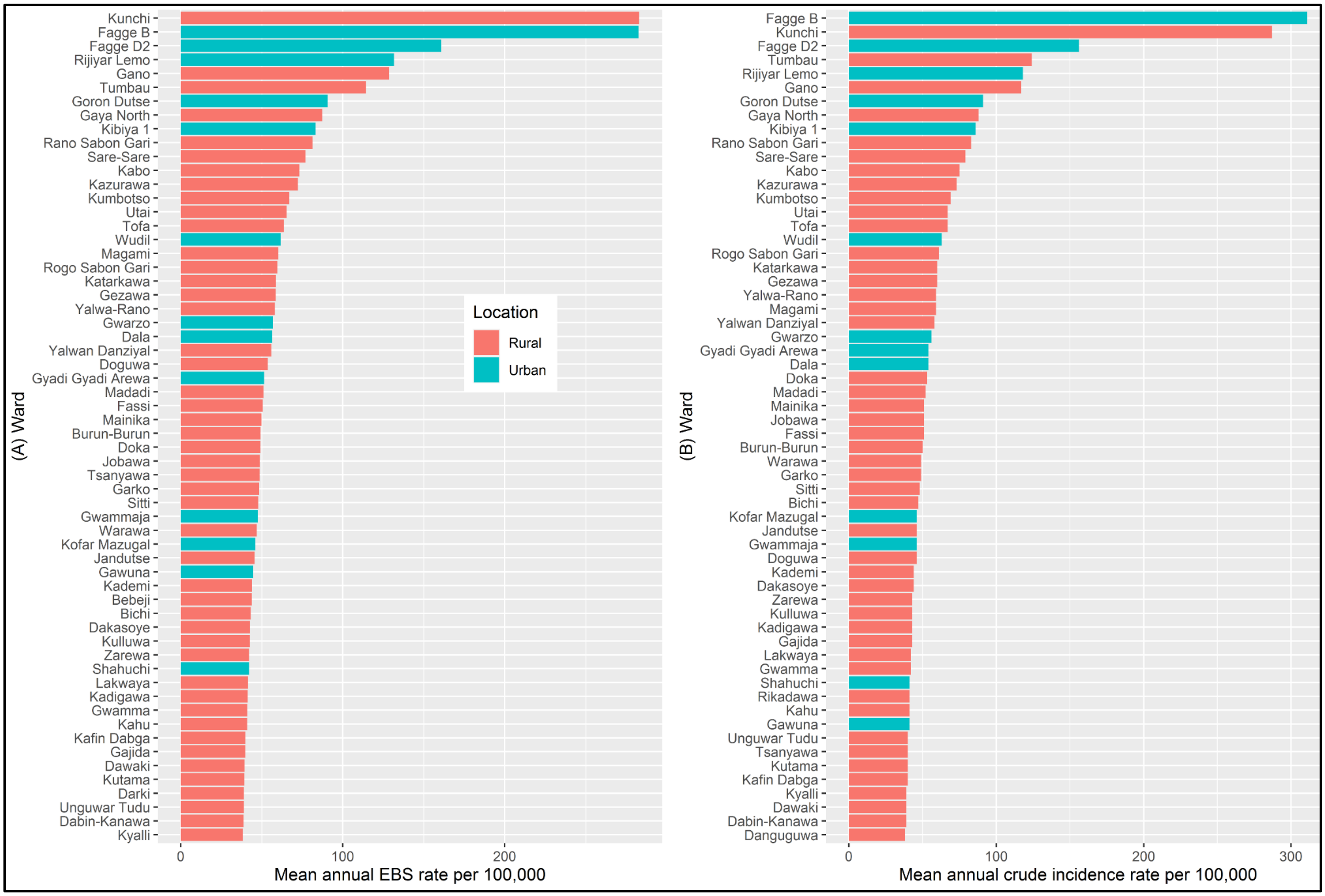


**S3 Fig. Distribution of mean annual EBS rate and mean annual crude incidence rate of cholera by urban and rural wards, 2010-2019.** Figure show (A) mean annual Empirical Bayes Smoothed (EBS) rates and (B) mean annual crude incidence rates per 100,000 for the top ranked 60 wards with heavy burden of cholera. (A) The top ranked ward with highest cholera rate is a rural ward (Kunchi) while (B) the top ranked ward with highest cholera burden is an urban ward (Fagge B). *Although there are differences between the EBS and crude rates, the same wards are represented; from top to bottom, only the respective positions of the wards on the graph changes.*

**S1 Table. Wards with the type of micro-hotspots by mean annual incidence rate and proportion of years with reported cholera cases (GTFCC method)**

| Type (T) of micro-hotspot | Ward | Location | Population | Mean Annual Incidence/100,000± | Proportion of years with reported cholera (%) |
|---|---|---|---|---|---|
| T1 | Fagge B | Urban | 6525 | 310.68 | 10.04 |
| T1 | Fagge D2 | Urban | 15539 | 156.03 | 10.47 |
| T1 | Rijiyar Lemo | Urban | 32200 | 117.56 | 14.10 |
| T1 | Gano | Rural | 38513 | 117.07 | 11.54 |
| T1 | Goron Dutse | Urban | 17259 | 90.51 | 9.40 |
| T1 | Gaya North | Rural | 46515 | 88.38 | 7.05 |
| T1 | Kibiya 1 | Rural | 15114 | 86.36 | 2.56 |
| T1 | Rano Sabon Gari | Rural | 27324 | 82.99 | 3.21 |
| T1 | Kabo | Rural | 27393 | 75.11 | 4.70 |
| T1 | Kumbotso | Urban | 15060 | 68.68 | 7.05 |
| T1 | Tofa | Rural | 19795 | 67.10 | 7.05 |
| T1 | Utai | Rural | 21892 | 67.02 | 3.42 |

| | | | | | |
|---|---|---|---|---|---|
| T1 | Wudil | Rural | 42977 | 62.52 | 6.84 |
| T1 | Rogo Sabon Gari | Rural | 28096 | 60.57 | 4.49 |
| T1 | Gezawa | Rural | 32091 | 59.69 | 7.05 |
| T1 | Gwarzo | Rural | 22648 | 56.40 | 6.20 |
| T1 | Dala | Urban | 41129 | 53.96 | 12.61 |
| T1 | Gyadi Gyadi Arewa | Urban | 8307 | 53.61 | 4.27 |
| T1 | Doka | Rural | 10652 | 53.18 | 3.21 |
| T1 | Madadi | Rural | 28477 | 51.93 | 3.42 |
| T1 | Mainika | Rural | 18458 | 51.31 | 4.06 |
| T1 | Warawa | Rural | 9087 | 49.39 | 3.21 |
| T1 | Garko | Rural | 33532 | 48.98 | 3.85 |
| T1 | Bichi | Rural | 73274 | 47.17 | 8.55 |
| T1 | Kofar Mazugal | Urban | 26356 | 46.10 | 9.83 |
| T1 | Jandutse | Rural | 41370 | 45.99 | 3.42 |
| T1 | Doguwa | Rural | 22303 | 45.56 | 3.85 |
| T1 | Gwammaja | Urban | 54069 | 45.55 | 12.61 |
| T1 | Kulluwa | Rural | 21976 | 43.40 | 3.21 |
| T1 | Gajida | Rural | 5569 | 43.36 | 2.56 |
| T1 | Lakwaya | Rural | 50658 | 41.80 | 5.34 |
| T1 | Gawuna | Urban | 31306 | 41.45 | 7.26 |
| T1 | Shahuchi | Urban | 40050 | 40.82 | 11.32 |
| T1 | Kafin Dabga | Rural | 24184 | 40.42 | 3.21 |
| T1 | Tsanyawa | Rural | 25466 | 40.22 | 4.06 |
| T1 | Kutama | Rural | 30525 | 39.95 | 4.06 |
| T1 | Unguwar Tudu | Rural | 25688 | 39.56 | 4.06 |
| T1 | Dawaki | Rural | 35337 | 39.08 | 6.84 |
| T1 | Danguguwa | Rural | 25931 | 38.26 | 2.99 |
| T1 | Rimin Gado | Rural | 20296 | 38.03 | 4.27 |
| T1 | Yanoko | Rural | 13048 | 37.48 | 3.42 |
| T1 | Sabon Gari-Wudil | Rural | 23346 | 36.01 | 4.91 |
| T1 | Tsakuwa | Rural | 29843 | 34.93 | 5.13 |
| T1 | Bebeji | Rural | 36646 | 34.05 | 3.85 |
| T1 | Ungogo | Urban | 37929 | 33.52 | 7.05 |
| T1 | Fagge A | Urban | 21523 | 33.47 | 3.85 |
| T1 | Bunkure | Rural | 35252 | 33.31 | 2.56 |
| T1 | Gwale | Urban | 34950 | 33.22 | 9.19 |
| T1 | Rijiyar Zaki | Rural | 40520 | 32.89 | 10.26 |
| T1 | Tsohon Gari | Rural | 29938 | 32.73 | 2.99 |
| T1 | Getso | Rural | 36029 | 32.17 | 3.63 |
| T1 | Tarauni | Urban | 12770 | 31.84 | 4.27 |
| T1 | Darki | Rural | 33384 | 30.83 | 3.21 |
| T1 | Dandago | Urban | 26536 | 29.53 | 7.26 |
| T1 | Zaitawa | Urban | 40975 | 28.61 | 9.83 |
| T1 | Sabon Gari | Rural | 30802 | 28.40 | 4.06 |
| T1 | Panshekara | Urban | 35135 | 28.31 | 6.20 |
| T1 | Rimin Dako | Rural | 24873 | 28.09 | 2.56 |
| T1 | Bagwai | Rural | 27981 | 28.01 | 3.63 |
| T1 | Kwajale | Rural | 20182 | 27.80 | 3.63 |
| T1 | She She | Urban | 22473 | 27.69 | 5.13 |
| T1 | Yalwa-Dala | Urban | 23474 | 27.49 | 4.91 |

| | | | | | |
|---|---|---|---|---|---|
| T1 | Kubaraci | Rural | 26001 | 27.21 | 2.99 |
| T1 | Yakasai | Urban | 36857 | 26.64 | 7.91 |
| T1 | Kiru | Rural | 53125 | 26.60 | 3.63 |
| T1 | Jakara | Urban | 22634 | 26.34 | 4.70 |
| T1 | Takai | Rural | 45279 | 26.05 | 3.85 |
| T1 | Sabon Gari West | Urban | 42804 | 25.81 | 5.77 |
| T1 | Shuwaki | Rural | 25746 | 25.44 | 3.42 |
| T1 | Danbare | Urban | 25292 | 25.41 | 3.85 |
| T1 | Dan Maliki | Urban | 106135 | 24.16 | 9.19 |
| T1 | Bachirawa | Urban | 73206 | 23.95 | 8.76 |
| T1 | Dakata | Urban | 49321 | 23.26 | 8.55 |
| T1 | Mandawari | Urban | 14175 | 22.06 | 3.42 |
| T1 | Lambu | Rural | 18732 | 21.97 | 2.78 |
| T1 | Gayawa | Rural | 50228 | 21.09 | 3.85 |
| T1 | Jalli | Rural | 21241 | 21.09 | 2.56 |
| T1 | Beli | Rural | 25641 | 20.49 | 2.78 |
| T1 | Bargoni | Rural | 34869 | 20.45 | 3.63 |
| T1 | Gyaranya | Urban | 17198 | 19.98 | 3.21 |
| T1 | Tanburawa | Rural | 33930 | 19.83 | 2.78 |
| T1 | Sani Mai Nagge | Urban | 41337 | 19.77 | 6.84 |
| T1 | Galadanchi | Urban | 24944 | 19.72 | 4.49 |
| T1 | Naibawa | Urban | 76213 | 19.65 | 6.62 |
| T1 | Yaryasa | Rural | 33669 | 19.65 | 2.56 |
| T1 | Chedi | Urban | 15050 | 19.40 | 2.78 |
| T1 | Dawakiji | Rural | 34966 | 19.04 | 3.21 |
| T1 | Dan Agundi | Urban | 18859 | 18.87 | 3.63 |
| T1 | Madobi | Rural | 29235 | 18.64 | 4.49 |
| T1 | Ketawa | Rural | 42522 | 18.20 | 3.63 |
| T1 | Tudun Fulani | Urban | 87983 | 18.08 | 6.20 |
| T1 | Sharada | Urban | 97279 | 18.03 | 10.68 |
| T1 | Ajingi | Rural | 25862 | 17.58 | 3.63 |
| T1 | Kara | Rural | 32141 | 17.21 | 3.21 |
| T1 | Kaura Goje | Urban | 101993 | 16.87 | 10.26 |
| T1 | Rogo Ruma | Rural | 32896 | 15.85 | 2.56 |
| T1 | Minjibir | Rural | 31563 | 15.71 | 2.78 |
| T1 | Kawaji | Urban | 119986 | 15.54 | 11.32 |
| T1 | Adakawa | Urban | 27219 | 15.23 | 4.06 |
| T1 | Gandu Albasa | Urban | 74484 | 14.55 | 5.34 |
| T1 | Kantudu | Urban | 26646 | 13.12 | 2.78 |
| T1 | Garun Gawa | Rural | 26221 | 13.10 | 4.27 |
| T1 | Kan Karofi | Urban | 38874 | 12.51 | 4.91 |
| T1 | Dorayi | Urban | 230272 | 12.30 | 14.32 |
| T1 | Jita | Rural | 25798 | 12.09 | 2.56 |
| T1 | Darmanawa | Urban | 67397 | 12.01 | 7.05 |
| T1 | Tudun Murtala | Urban | 83078 | 11.62 | 7.69 |
| T1 | Zango-III | Urban | 64141 | 11.51 | 7.26 |
| T1 | Kausani | Rural | 32945 | 11.19 | 2.99 |
| T1 | Kofar Ruwa | Urban | 101663 | 11.12 | 8.76 |
| T1 | Dankaza | Rural | 29476 | 10.88 | 3.21 |
| T1 | Gobirawa | Urban | 222097 | 10.71 | 11.11 |

| | | | | | |
|---|---|---|---|---|---|
| T1 | Hotoro South | Urban | 59253 | 10.62 | 5.13 |
| T1 | Gabasawa | Rural | 44889 | 10.48 | 2.78 |
| T1 | Gwangwan | Rural | 31455 | 10.44 | 2.99 |
| **T1 Total** | **115** | | **4,505,370** | | |
| T2 | Kunchi | Rural | 16152 | 287.17 | 1.50 |
| T2 | Tumbau | Rural | 25985 | 123.76 | 2.35 |
| T2 | Sare-Sare | Rural | 21673 | 78.77 | 1.07 |
| T2 | Kazurawa | Rural | 35668 | 73.13 | 1.71 |
| T2 | Katarkawa | Rural | 13218 | 59.61 | 0.85 |
| T2 | Yalwa-Rano | Rural | 24823 | 59.19 | 1.92 |
| T2 | Magami | Rural | 56336 | 59.18 | 1.50 |
| T2 | Yalwan Danziyal | Rural | 16232 | 57.69 | 1.71 |
| T2 | Jobawa | Rural | 15728 | 51.36 | 0.64 |
| T2 | Fassi | Rural | 35407 | 51.19 | 0.85 |
| T2 | Burun-Burun | Rural | 29759 | 49.87 | 2.14 |
| T2 | Sitti | Rural | 44455 | 48.25 | 2.35 |
| T2 | Kademi | Rural | 38155 | 44.44 | 1.28 |
| T2 | Dakasoye | Rural | 17303 | 44.11 | 1.92 |
| T2 | Kadigawa | Rural | 10735 | 42.82 | 0.64 |
| T2 | Zarewa | Rural | 28438 | 42.80 | 1.50 |
| T2 | Gwamma | Rural | 24776 | 41.76 | 1.92 |
| T2 | Kahu | Rural | 26805 | 41.46 | 1.07 |
| T2 | Rikadawa | Rural | 19416 | 41.00 | 1.92 |
| T2 | Dabin-Kanawa | Rural | 24778 | 39.22 | 1.07 |
| T2 | Kyalli | Rural | 35928 | 38.90 | 1.71 |
| T2 | Gurjiya-I | Rural | 18573 | 35.27 | 1.28 |
| T2 | Yako | Rural | 22236 | 35.25 | 1.07 |
| T2 | Mesar Tudu | Rural | 27990 | 35.19 | 1.71 |
| T2 | Zurgu | Rural | 22851 | 34.63 | 2.14 |
| T2 | Yammedi | Rural | 20273 | 34.34 | 1.92 |
| T2 | Durba | Rural | 13432 | 33.21 | 0.85 |
| T2 | Yalwa Karama | Rural | 5348 | 32.97 | 2.35 |
| T2 | Jili | Rural | 12182 | 30.77 | 1.50 |
| T2 | Dindere | Rural | 5710 | 30.08 | 2.35 |
| T2 | Wudilawa | Rural | 15880 | 29.39 | 1.50 |
| T2 | Sumaila | Rural | 35188 | 29.14 | 2.14 |
| T2 | Kumurya | Rural | 36284 | 28.79 | 1.92 |
| T2 | Mekiya | Rural | 34634 | 27.86 | 0.64 |
| T2 | Yada Kwari | Rural | 20308 | 27.81 | 0.21 |
| T2 | Lausu | Rural | 23904 | 26.18 | 2.14 |
| T2 | Magajin Gari | Rural | 10462 | 25.67 | 1.92 |
| T2 | Shanono | Rural | 33914 | 25.02 | 2.35 |
| T2 | Sabon Birni | Rural | 21624 | 24.90 | 2.35 |
| T2 | Kadawa | Rural | 17158 | 24.87 | 0.85 |
| T2 | Madachi | Rural | 17583 | 24.46 | 1.92 |
| T2 | Achika | Rural | 13973 | 24.44 | 2.35 |
| T2 | Langel | Rural | 10551 | 23.87 | 1.71 |
| T2 | Gani | Rural | 37915 | 23.57 | 0.64 |
| T2 | Tsaure | Rural | 29476 | 23.50 | 0.85 |

| | | | | | |
|---|---|---|---|---|---|
| T2 | Dal | Rural | 34797 | 23.40 | 0.85 |
| T2 | Unguwar Gai | Rural | 23092 | 23.25 | 0.64 |
| T2 | Duja | Rural | 10357 | 22.82 | 0.64 |
| T2 | Kafin Malamai | Rural | 26812 | 21.55 | 1.50 |
| T2 | Kogo | Rural | 21341 | 21.29 | 1.92 |
| T2 | Dadin Kowa | Rural | 36464 | 20.74 | 2.14 |
| T2 | Burji | Rural | 10180 | 20.64 | 1.07 |
| T2 | Kwamarawa | Rural | 22976 | 20.53 | 1.28 |
| T2 | Gulu | Rural | 18687 | 20.29 | 1.92 |
| T2 | Yan Dala | Rural | 8547 | 20.03 | 1.07 |
| T2 | Chirin | Rural | 24971 | 19.98 | 1.28 |
| T2 | Zinyau | Rural | 13007 | 19.71 | 1.71 |
| T2 | Karfi | Rural | 19765 | 19.32 | 1.07 |
| T2 | Albasu Central | Rural | 33373 | 18.89 | 1.71 |
| T2 | Cinkoso | Rural | 6055 | 18.71 | 1.50 |
| T2 | Saya-Saya | Rural | 33325 | 18.54 | 1.71 |
| T2 | Imawa | Rural | 11918 | 17.94 | 0.43 |
| T2 | Dalawa | Rural | 27934 | 17.87 | 1.07 |
| T2 | Barkum | Rural | 24521 | 17.86 | 2.14 |
| T2 | Kunkurawa | Rural | 25956 | 17.71 | 0.64 |
| T2 | Gurjiya-III | Rural | 24342 | 17.27 | 1.07 |
| T2 | Rumo | Rural | 16793 | 17.20 | 0.43 |
| T2 | Farun Ruwa | Rural | 40563 | 17.11 | 0.64 |
| T2 | Rahama | Rural | 23350 | 17.09 | 1.50 |
| T2 | Saidawa | Rural | 35860 | 17.01 | 0.64 |
| T2 | Daho | Rural | 24652 | 16.92 | 1.07 |
| T2 | Fanda | Rural | 32148 | 16.34 | 0.85 |
| T2 | Shagogo | Rural | 32505 | 16.34 | 1.71 |
| T2 | Yankatsari | Rural | 10589 | 16.26 | 1.92 |
| T2 | Fammar | Rural | 22711 | 16.19 | 0.43 |
| T2 | Danzabuwa | Rural | 53451 | 15.91 | 1.50 |
| T2 | Unguwar Tsohuwa | Rural | 16645 | 15.78 | 1.28 |
| T2 | Gaya South | Rural | 40232 | 15.60 | 2.35 |
| T2 | Janguza | Rural | 11936 | 15.45 | 2.14 |
| T2 | Saji | Rural | 21225 | 15.24 | 1.71 |
| T2 | Diso | Urban | 9842 | 14.83 | 1.92 |
| T2 | Fagwalawa | Rural | 31663 | 14.74 | 1.28 |
| T2 | Kosawa | Rural | 24617 | 14.74 | 0.85 |
| T2 | Gafasa | Rural | 25891 | 14.71 | 0.64 |
| T2 | Unguwar Rimi-Tofa | Rural | 12554 | 13.84 | 1.28 |
| T2 | Kuki | Rural | 18341 | 13.78 | 1.28 |
| T2 | Karfi-Kura | Rural | 30723 | 13.74 | 1.71 |
| T2 | Zainabi | Rural | 15045 | 13.56 | 0.85 |
| T2 | Tagwaye | Rural | 21696 | 13.30 | 1.50 |
| T2 | Balare | Rural | 24819 | 13.19 | 0.85 |
| T2 | Nariya | Rural | 21249 | 12.98 | 1.50 |
| T2 | Yarimawa | Rural | 9178 | 12.84 | 1.71 |
| T2 | Jauben Kudu | Rural | 7153 | 12.32 | 1.28 |
| T2 | Kauran Mata | Rural | 13039 | 12.30 | 1.71 |
| T2 | Dosan | Rural | 25289 | 11.23 | 1.50 |

| | | | | | |
|---|---|---|---|---|---|
| T2 | Gafan | Rural | 27261 | 11.05 | 1.28 |
| T2 | Yankamaye | Rural | 29327 | 11.02 | 1.07 |
| T2 | Gora | Rural | 27969 | 10.80 | 1.92 |
| T2 | Danlasan | Rural | 6123 | 10.73 | 0.43 |
| T2 | Tudun Kaya | Rural | 27046 | 10.71 | 1.28 |
| T2 | Yargaya | Rural | 19496 | 10.61 | 0.85 |
| T2 | Zuwo | Rural | 22652 | 10.51 | 1.07 |
| T2 | Indabo | Rural | 15059 | 10.46 | 1.71 |
| T2 | Butu-Butu | Rural | 16450 | 10.18 | 1.28 |
| T2 | Rano Dawaki | Rural | 20510 | 10.00 | 1.07 |
| **T2 Total** | **105** | | **2,413,291** | | |
| T3 | Unguwa Uku Cikin Gari | Urban | 35175 | 9.96 | 2.56 |
| T3 | Bakin Ruwa | Urban | 37500 | 9.94 | 4.49 |
| T3 | Kadawa-Ungogo | Urban | 56668 | 9.62 | 3.42 |
| T3 | Tudun Wazurchi | Urban | 52243 | 9.40 | 4.49 |
| T3 | Jogana | Rural | 52963 | 9.19 | 4.27 |
| T3 | Madigawa | Urban | 21297 | 8.78 | 2.99 |
| T3 | Gwagwarwa | Urban | 56368 | 8.68 | 3.21 |
| T3 | Fanisau | Rural | 27616 | 8.67 | 2.56 |
| T3 | Zango-IV | Urban | 112963 | 8.56 | 5.98 |
| T3 | Zoza | Rural | 44385 | 8.50 | 2.99 |
| T3 | Kabuga | Urban | 135672 | 8.17 | 9.19 |
| T3 | Dagumawa | Rural | 39250 | 8.00 | 3.21 |
| T3 | Mariri | Urban | 49866 | 7.92 | 3.21 |
| T3 | Unguwa Uku Kauyen Alu | Urban | 64103 | 7.26 | 4.27 |
| T3 | Kwachiri | Urban | 94577 | 6.47 | 3.85 |
| T3 | Zango-I | Rural | 48551 | 5.93 | 2.78 |
| T3 | Hotoro | Urban | 58150 | 5.68 | 2.78 |
| T3 | Giginyu | Urban | 163524 | 5.59 | 6.84 |
| T3 | Gama | Urban | 79001 | 5.05 | 4.06 |
| T3 | Badume | Rural | 56171 | 4.80 | 2.56 |
| T3 | Rangaza | Rural | 40089 | 4.66 | 2.78 |
| T3 | Hotoro North | Urban | 142720 | 4.46 | 5.13 |
| T3 | Dawanau | Rural | 101627 | 2.26 | 2.78 |
| **T3 Total** | **23** | | **1,570,479** | | |
| T4 | Dogon Kawo | Rural | 17125 | 9.97 | 0.43 |
| T4 | Dawaki West | Rural | 18744 | 9.92 | 1.92 |
| T4 | Rimi | Rural | 33877 | 9.69 | 0.21 |
| T4 | Falgore Doguwa | Rural | 23702 | 9.43 | 1.71 |
| T4 | Tumfafi | Rural | 57525 | 9.37 | 1.50 |
| T4 | Dawaki East | Rural | 21121 | 9.33 | 0.21 |
| T4 | Dalili | Rural | 18393 | 9.28 | 1.28 |
| T4 | Jama'A | Rural | 14354 | 9.26 | 1.28 |
| T4 | Tanagar | Rural | 26368 | 9.21 | 0.85 |
| T4 | Karefa | Rural | 25742 | 9.12 | 1.07 |
| T4 | Kwas | Rural | 18859 | 9.05 | 0.64 |
| T4 | Zango-II | Rural | 14828 | 8.64 | 1.50 |
| T4 | Amarawa | Rural | 6311 | 8.61 | 0.64 |
| T4 | Yan Barau | Rural | 10892 | 8.44 | 1.28 |
| T4 | Goron Maje | Rural | 18340 | 8.19 | 0.43 |

| | | | | | |
|---|---|---|---|---|---|
| T4 | Tudun Wada | Urban | 23443 | 8.08 | 1.71 |
| T4 | Gawo | Rural | 30519 | 8.06 | 2.35 |
| T4 | Sararin Gezawa | Rural | 24744 | 7.96 | 1.71 |
| T4 | Maraku | Rural | 34386 | 7.82 | 1.07 |
| T4 | Kwankwaso | Rural | 27025 | 7.71 | 2.35 |
| T4 | Danbagina | Rural | 25664 | 7.67 | 0.43 |
| T4 | Daura | Rural | 23781 | 7.50 | 0.64 |
| T4 | Bauda | Rural | 20891 | 7.46 | 1.07 |
| T4 | Daurawa | Urban | 15800 | 7.46 | 1.50 |
| T4 | Waire | Rural | 25261 | 7.43 | 1.07 |
| T4 | Turawa | Rural | 21952 | 7.36 | 1.50 |
| T4 | Yalwa-Kiru | Rural | 48008 | 7.25 | 1.07 |
| T4 | Rurum Tsohon Gari | Rural | 21589 | 7.07 | 1.07 |
| T4 | Garun Malam | Rural | 15330 | 6.91 | 1.07 |
| T4 | Ginsawa | Rural | 5511 | 6.91 | 0.64 |
| T4 | Riruwai | Rural | 23051 | 6.84 | 1.07 |
| T4 | Faragai | Rural | 29036 | 6.78 | 1.07 |
| T4 | Kafin Agur | Rural | 9952 | 6.69 | 1.07 |
| T4 | Dawakin Gulu | Rural | 5967 | 6.60 | 0.43 |
| T4 | Gadanya | Rural | 24723 | 6.37 | 0.43 |
| T4 | Fulatan | Rural | 30147 | 5.99 | 1.07 |
| T4 | Garin Dau | Rural | 17685 | 5.94 | 1.07 |
| T4 | Zogarawa | Rural | 10913 | 5.91 | 1.07 |
| T4 | Shamakawa | Rural | 11706 | 5.89 | 0.64 |
| T4 | Jido | Rural | 15637 | 5.88 | 0.85 |
| T4 | Gammo | Rural | 20742 | 5.87 | 0.64 |
| T4 | Rurum Sabon Gari | Rural | 28995 | 5.86 | 1.71 |
| T4 | Kabuwaya | Urban | 16600 | 5.84 | 1.5 |
| T4 | Godiya | Rural | 30333 | 5.79 | 0.64 |
| T4 | Dambatta West | Rural | 32011 | 5.79 | 1.5 |
| T4 | Babban Giji | Urban | 39643 | 5.74 | 2.14 |
| T4 | Yallami | Rural | 32776 | 5.73 | 1.5 |
| T4 | Zuga | Rural | 26023 | 5.68 | 1.28 |
| T4 | Katumari | Rural | 16426 | 5.60 | 0.21 |
| T4 | Sakara Tsa | Rural | 9430 | 5.57 | 0.21 |
| T4 | Tudun Nufawa | Urban | 33552 | 5.55 | 2.35 |
| T4 | Gwarmai-Bebeji | Rural | 51710 | 5.40 | 1.5 |
| T4 | Lajawa | Rural | 30135 | 5.33 | 1.5 |
| T4 | Dangora | Rural | 21748 | 5.28 | 0.64 |
| T4 | Garun Sheme | Rural | 8739 | 5.26 | 0.43 |
| T4 | Kau-Kau | Rural | 25594 | 5.24 | 1.92 |
| T4 | Makoda | Rural | 36051 | 5.15 | 0.64 |
| T4 | Magajin Hajji | Rural | 23602 | 5.12 | 0.64 |
| T4 | Jajaye | Rural | 35409 | 5.06 | 1.07 |
| T4 | Sanda | Rural | 23000 | 5.06 | 0.85 |
| T4 | Koguna | Rural | 31300 | 5.04 | 0.85 |
| T4 | Sarina | Rural | 26202 | 5.01 | 0.43 |
| T4 | Yammata | Urban | 34716 | 5.01 | 0.85 |
| T4 | Tarai | Rural | 28632 | 4.99 | 1.07 |
| T4 | Rantan | Rural | 16188 | 4.97 | 0.43 |

| | | | | | |
|---|---|---|---|---|---|
| T4 | Baburi | Rural | 39019 | 4.71 | 1.07 |
| T4 | Jibga | Rural | 47908 | 4.62 | 0.64 |
| T4 | Nata-Ala | Rural | 43657 | 4.47 | 1.71 |
| T4 | Wangara | Rural | 32803 | 4.34 | 1.92 |
| T4 | Babbar Ruga | Rural | 34707 | 4.22 | 0.64 |
| T4 | Durbunde | Rural | 34308 | 4.17 | 0.85 |
| T4 | Tamawa | Rural | 12671 | 4.15 | 0.43 |
| T4 | Yada Kunya | Rural | 25440 | 4.06 | 1.07 |
| T4 | Wangara-Tofa | Rural | 9593 | 4.06 | 0.43 |
| T4 | Kantama | Rural | 31602 | 4.05 | 0.64 |
| T4 | Dukawa | Rural | 28412 | 4.02 | 1.5 |
| T4 | Kuka | Rural | 22546 | 4.00 | 0.85 |
| T4 | Romo | Rural | 26268 | 4.00 | 1.07 |
| T4 | Gurjiya-II | Rural | 12807 | 3.97 | 0.85 |
| T4 | Kiyawa | Rural | 30620 | 3.78 | 0.64 |
| T4 | Zarogi | Rural | 23641 | 3.66 | 0.43 |
| T4 | Gogel | Rural | 7495 | 3.50 | 0.43 |
| T4 | Kwanyawa | Rural | 27754 | 3.48 | 0.64 |
| T4 | Alajawa | Rural | 15173 | 3.46 | 0.43 |
| T4 | Yanganau | Rural | 28795 | 3.43 | 0.64 |
| T4 | Raba | Rural | 26802 | 3.43 | 0.64 |
| T4 | Rigar Duka | Rural | 19089 | 3.42 | 1.07 |
| T4 | Garo | Rural | 43276 | 3.30 | 1.07 |
| T4 | Tsaudawa | Rural | 17191 | 3.22 | 0.64 |
| T4 | Gamarya | Rural | 20303 | 3.19 | 0.85 |
| T4 | Chiranchi | Urban | 63634 | 3.11 | 2.14 |
| T4 | Kwami | Rural | 3724 | 3.08 | 0.21 |
| T4 | Gargari | Rural | 32536 | 3.02 | 1.5 |
| T4 | Gundutse | Rural | 17151 | 2.97 | 0.64 |
| T4 | Garun Danga | Rural | 31341 | 2.93 | 0.64 |
| T4 | Muntsira | Rural | 40553 | 2.91 | 1.5 |
| T4 | Shuwaki-Kunchi | Rural | 24081 | 2.88 | 0.43 |
| T4 | Maraku-Kiru | Rural | 16707 | 2.85 | 0.64 |
| T4 | Dangada | Rural | 18701 | 2.81 | 0.43 |
| T4 | Daddarawa | Rural | 14007 | 2.63 | 0.43 |
| T4 | Ruwan Bago | Rural | 56456 | 2.56 | 1.28 |
| T4 | Joda | Rural | 31624 | 2.54 | 0.43 |
| T4 | Babawa | Rural | 51716 | 2.53 | 1.71 |
| T4 | Kunya | Rural | 46705 | 2.48 | 0.64 |
| T4 | Sarkin Kura | Rural | 19914 | 2.47 | 0.64 |
| T4 | Chiromawa | Rural | 24043 | 2.46 | 0.64 |
| T4 | Dambatta East | Rural | 47500 | 2.37 | 0.85 |
| T4 | Dugurawa | Rural | 5580 | 2.35 | 0.21 |
| T4 | Ajumawa | Rural | 28034 | 2.34 | 0.43 |
| T4 | Saye | Rural | 22125 | 2.15 | 0.85 |
| T4 | Wasai | Rural | 30178 | 2.08 | 0.64 |
| T4 | Doka Dawa | Rural | 12920 | 2.03 | 0.43 |
| T4 | San San | Rural | 19029 | 2.00 | 0.43 |
| T4 | Fagge D1 | Urban | 5981 | 1.99 | 0.21 |
| T4 | Falali | Rural | 25657 | 1.98 | 0.85 |

| | | | | | |
|---|---|---|---|---|---|
| T4 | Shere | Rural | 20152 | 1.96 | 0.43 |
| T4 | Dan Hassan | Rural | 31836 | 1.91 | 0.85 |
| T4 | Sabon Gari East | Urban | 39356 | 1.84 | 0.64 |
| T4 | Fajewa | Rural | 21549 | 1.83 | 0.64 |
| T4 | Kurugu | Rural | 21147 | 1.80 | 0.64 |
| T4 | Unguwar Gano | Urban | 16183 | 1.79 | 0.43 |
| T4 | Shakogi | Rural | 22562 | 1.77 | 0.64 |
| T4 | Jemagu | Rural | 7451 | 1.76 | 0.21 |
| T4 | Falgore Rogo | Rural | 34411 | 1.73 | 0.21 |
| T4 | Karofin Yashi | Rural | 14574 | 1.69 | 0.43 |
| T4 | Kureken Sani | Rural | 24439 | 1.69 | 0.64 |
| T4 | Gamoji | Rural | 14666 | 1.68 | 0.43 |
| T4 | Yautar Kudu | Rural | 39144 | 1.60 | 0.64 |
| T4 | Yola | Rural | 15463 | 1.59 | 0.43 |
| T4 | Baawa | Rural | 37046 | 1.55 | 0.85 |
| T4 | Zakarawa | Rural | 18207 | 1.44 | 0.43 |
| T4 | Yakun | Rural | 9345 | 1.41 | 0.21 |
| T4 | Durmawa | Rural | 18862 | 1.39 | 0.21 |
| T4 | Gala | Rural | 29479 | 1.34 | 0.43 |
| T4 | Karo | Rural | 11129 | 1.18 | 0.21 |
| T4 | Jigawa | Rural | 22985 | 1.16 | 0.43 |
| T4 | Masu | Rural | 45507 | 1.15 | 0.43 |
| T4 | Unguwar Duniya | Rural | 11429 | 1.15 | 0.21 |
| T4 | Wailare | Rural | 23158 | 1.13 | 0.43 |
| T4 | Tattarawa | Rural | 22547 | 1.13 | 0.43 |
| T4 | Chalawa | Rural | 12884 | 1.13 | 0.21 |
| T4 | Kibiya 2 | Rural | 11363 | 1.12 | 0.21 |
| T4 | Tsamiya Babba | Rural | 60305 | 1.11 | 0.85 |
| T4 | Bagwaro | Rural | 24433 | 1.07 | 0.43 |
| T4 | Marke | Rural | 34959 | 1.02 | 0.64 |
| T4 | Gediya | Rural | 39586 | 1.00 | 0.64 |
| T4 | Hawaden Galadima | Rural | 12466 | 0.99 | 0.21 |
| T4 | Dogon Nama | Urban | 40635 | 0.98 | 0.64 |
| T4 | Balan | Rural | 26642 | 0.95 | 0.21 |
| T4 | Dugabau | Rural | 28064 | 0.94 | 0.43 |
| T4 | Gwarabjawa | Rural | 46165 | 0.93 | 0.43 |
| T4 | Wak | Rural | 12331 | 0.93 | 0.21 |
| T4 | Wuro Bagga | Rural | 28044 | 0.91 | 0.43 |
| T4 | Ganduje | Rural | 16053 | 0.90 | 0.21 |
| T4 | Faruruwa | Rural | 15059 | 0.90 | 0.21 |
| T4 | Azore | Rural | 28478 | 0.83 | 0.21 |
| T4 | Gargai | Rural | 14177 | 0.81 | 0.21 |
| T4 | Dashi | Rural | 16334 | 0.80 | 0.21 |
| T4 | Dunbulum | Rural | 18525 | 0.78 | 0.21 |
| T4 | Bataiya | Rural | 33718 | 0.78 | 0.43 |
| T4 | Masanawa | Rural | 15791 | 0.78 | 0.21 |
| T4 | Chula | Rural | 16919 | 0.78 | 0.21 |
| T4 | Galadimawa | Rural | 17352 | 0.76 | 0.21 |
| T4 | Toranke | Rural | 52242 | 0.75 | 0.43 |
| T4 | Badafi | Rural | 36855 | 0.71 | 0.43 |

| | | | | | |
|---|---|---|---|---|---|
| T4 | Garun Babba | Rural | 19425 | 0.70 | 0.21 |
| T4 | Tariwa | Rural | 17349 | 0.66 | 0.21 |
| T4 | Gyadi Gyadi Kudu | Urban | 20081 | 0.65 | 0.21 |
| T4 | Tangaji | Rural | 20975 | 0.63 | 0.21 |
| T4 | Dundun | Rural | 21302 | 0.62 | 0.21 |
| T4 | Durun | Rural | 21579 | 0.61 | 0.21 |
| T4 | Kofa | Rural | 18876 | 0.61 | 0.21 |
| T4 | Garin Ali | Rural | 21853 | 0.60 | 0.21 |
| T4 | Makwaro | Rural | 25221 | 0.58 | 0.21 |
| T4 | Kanwa | Rural | 23119 | 0.57 | 0.21 |
| T4 | Tabo | Rural | 23680 | 0.55 | 0.21 |
| T4 | Yandadi | Rural | 24158 | 0.54 | 0.21 |
| T4 | Tatsan | Rural | 28379 | 0.53 | 0.21 |
| T4 | Kadan Dani | Rural | 49980 | 0.53 | 0.21 |
| T4 | Ranka | Rural | 22144 | 0.52 | 0.21 |
| T4 | Hungu | Rural | 22528 | 0.51 | 0.21 |
| T4 | Gandurwawa | Rural | 24932 | 0.49 | 0.21 |
| T4 | Dabar Kwari | Rural | 28440 | 0.46 | 0.21 |
| T4 | Kwa | Rural | 25755 | 0.46 | 0.21 |
| T4 | Gogori | Rural | 25295 | 0.45 | 0.21 |
| T4 | Gurun | Rural | 30625 | 0.43 | 0.21 |
| T4 | Kwarkiya | Rural | 30110 | 0.41 | 0.21 |
| T4 | Dutsen Bakoshi | Rural | 34245 | 0.37 | 0.21 |
| T4 | Tsakuwa-Minjibir | Rural | 35157 | 0.35 | 0.21 |
| T4 | Kachako | Rural | 49083 | 0.27 | 0.21 |
| T4 | Anadariya | Rural | 9164 | 0.00 | 0 |
| T4 | Baguda | Rural | 13766 | 0.00 | 0 |
| T4 | Bono | Rural | 24015 | 0.00 | 0 |
| T4 | Bumai | Rural | 13945 | 0.00 | 0 |
| T4 | Damau | Rural | 15128 | 0.00 | 0 |
| T4 | Dansoshiya | Rural | 17467 | 0.00 | 0 |
| T4 | Dorawar Sallau | Rural | 14148 | 0.00 | 0 |
| T4 | Durma | Rural | 16360 | 0.00 | 0 |
| T4 | Fagge C | Urban | 10015 | 0.00 | 0 |
| T4 | Fagolo | Rural | 23647 | 0.00 | 0 |
| T4 | Fankurun | Rural | 9004 | 0.00 | 0 |
| T4 | Gagarame | Rural | 39982 | 0.00 | 0 |
| T4 | Galinja | Rural | 7031 | 0.00 | 0 |
| T4 | Garfa | Rural | 23711 | 0.00 | 0 |
| T4 | Goron-Dutse | Rural | 19404 | 0.00 | 0 |
| T4 | Gozarki | Rural | 13289 | 0.00 | 0 |
| T4 | Gude | Rural | 19450 | 0.00 | 0 |
| T4 | Gurduba | Rural | 23870 | 0.00 | 0 |
| T4 | Gwanda | Rural | 28518 | 0.00 | 0 |
| T4 | Gwarmai-Kunchi | Rural | 19109 | 0.00 | 0 |
| T4 | Juma Galadima | Rural | 19989 | 0.00 | 0 |
| T4 | Kabagiwa | Rural | 28427 | 0.00 | 0 |
| T4 | Kadamu | Rural | 18850 | 0.00 | 0 |
| T4 | Kanawa | Rural | 24017 | 0.00 | 0 |
| T4 | Kanwa-Kabo | Rural | 15512 | 0.00 | 0 |

| | | | | | |
|---|---|---|---|---|---|
| T4 | Karmami | Rural | 35292 | 0.00 | 0 |
| T4 | Kasuwar Kuka | Rural | 24469 | 0.00 | 0 |
| T4 | Kokiya | Rural | 16483 | 0.00 | 0 |
| T4 | Kore | Rural | 30057 | 0.00 | 0 |
| T4 | Kuru | Rural | 32755 | 0.00 | 0 |
| T4 | Kurun Sumau | Rural | 20590 | 0.00 | 0 |
| T4 | Leni | Rural | 9277 | 0.00 | 0 |
| T4 | Madari | Rural | 14706 | 0.00 | 0 |
| T4 | Maimakawa | Rural | 36008 | 0.00 | 0 |
| T4 | Maitsidau | Rural | 30289 | 0.00 | 0 |
| T4 | Matan Fada | Rural | 19367 | 0.00 | 0 |
| T4 | Ridawa | Rural | 7590 | 0.00 | 0 |
| T4 | Sarbi | Rural | 14062 | 0.00 | 0 |
| T4 | Satame | Rural | 24776 | 0.00 | 0 |
| T4 | Tamburawan Gabas | Rural | 13850 | 0.00 | 0 |
| T4 | Tanawa | Urban | 9808 | 0.00 | 0 |
| T4 | Tarauni-Gabasawa | Rural | 15539 | 0.00 | 0 |
| T4 | Tsakiya | Rural | 20519 | 0.00 | 0 |
| T4 | Tsangaya | Rural | 30886 | 0.00 | 0 |
| T4 | Unguwar Bai | Rural | 23941 | 0.00 | 0 |
| T4 | Unguwar Rimi | Rural | 16526 | 0.00 | 0 |
| T4 | Yangizo | Rural | 10682 | 0.00 | 0 |
| T4 | Yautar Arewa | Rural | 20706 | 0.00 | 0 |
| T4 | Yumbu | Rural | 17721 | 0.00 | 0 |
| T4 | Zakirai | Rural | 24873 | 0.00 | 0 |
| T4 | Zugachi | Rural | 26122 | 0.00 | 0 |
| **T4 Total** | **241** | | **5,811,246** | | |

±Note: Table is arranged in descending order of mean annual incidence within the micro-hotspots types (T). The ward population reflects the year 2019 estimates. Out the 484 wards in Kano State, GTFCC method classified 115, 105, 23, and 241 as T1, T2, T3, and T4, respectively.
